# Preventing COVID-19 Outbreaks Through Surveillance Testing in Healthcare Facilities - A Modelling Study

**DOI:** 10.1101/2021.05.03.21255976

**Authors:** Tim Litwin, Jens Timmer, Mathias Berger, Andreas Wahl-Kordon, Matthias J. Müller, Clemens Kreutz

## Abstract

Surveillance testing within healthcare facilities provides an opportunity to prevent severe outbreaks of coronavirus disease 2019 (COVID-19). However, the quantitative impact of different available surveillance strategies is not well-understood. Our study adds to the available body of evidence by examining different strategies for their potential to decrease the probability of outbreaks in these facilities. Based on our findings, we propose determinants of successful surveillance measures. To this end, we establish an individual-based model representative of a mental health hospital yielding generalizable results. Attributes and features of this facility were derived from a prototypical hospital, which provides psychiatric, psychosomatic and psychotherapeutic treatment. We estimate the relative reduction of outbreak probability for three test strategies (entry test, once-weekly test and twice-weekly test) relative to a symptom-based baseline strategy. We found that fast diagnostic test results and adequate compliance of the clinic population are mandatory for conducting effective surveillance. The robustness of these results towards uncertainties is demonstrated via comprehensive sensitivity analyses. In summary, we robustly quantified the efficacy of different surveillance scenarios and conclude that active testing in mental health hospitals and similar facilities successfully reduces the number of COVID-19 outbreaks.

## Introduction

Treatment and care facilities with intermediate to long-term treatment durations pose a setting in which active COVID-19 surveillance strategies are urgently required. This became apparent from the numerous reports of disastrous outbreaks in skilled nursing facilities [1, 2] which host a population at an age-related high risk of fatal disease courses [3]. Psychiatric and psychosomatic facilities are faced with the challenge to maintain and ensure patient and staff security with at the same time increasing psychiatric symptoms in patients [4] and safety concerns in regard to a possible hospital stay. Establishing ways to effectively protect these populations allows these facilities to continue their regular functions despite of the current circumstances. Thus, these facilities in particular may substantially benefit from interventions intended to decrease the risk of possible COVID-19 outbreaks.

Surveillance aims at the disruption of evolving infection clusters in order to halt the spread of infection. Existing COVID-19 surveillance strategies revolve around detection of infected individuals in order to isolate them and their close contacts. To this end, possible infections are commonly confirmed by polymerase chain reaction (PCR) tests [5]. Yet, such tests are subject to certain structural constraints as they need to be performed in a laboratory which effectively delays the initiation of isolation and tracing. To circumvent these constraints, the use of point-of-care (PoC) tests in the form of faster but less sensitive antigen tests has been proposed. Although those tests might only detect individuals with high enough viral loads to actually be infectious [6], the fact that those tests can be carried out by each individual autonomously allows to make at least a tentative COVID-19 diagnosis with greater flexibility and in a less centralized manner. This highlights their possible use for surveillance in single institutions.

The efficacy of surveillance testing in preventing outbreaks can be either assessed by empirical or by more conceptual means. Empirical observational studies are a necessary tool to validate the benefits of implementing surveillance. However, these studies are difficult to conduct correctly since the heterogeneity of treatment facilities and settings impairs the comparability of the impact of different interventions. Hence, modelling studies are essential to complement empirical evidence. They are considerably less restrictive in terms of possible scientific questions that can be answered and allow for the investigation of interventions under clearly specified assumptions. Modelling allows to conceptualize the important aspects of the problem [7] and to identify the main determinants responsible for observed phenomena.

High quality evidence about the benefits of surveillance testing is sparse. However, there are existing modelling studies that tackle different aspects of surveillance testing, such as limited availability of test resources [8], cohorting of staff and residents [9], and regular testing of staff/residents at different frequencies [10]. Those results are of limited generalizability: Most models report outcomes for which a reasonable quantification of uncertainties is impossible, e.g. the cumulative number of infections at a late stage of the outbreak. Uncertainties about the structure of the infection spread and the underlying epidemiological parameters amplify the uncertainties of the outcome in a non-linear manner which complicates the conduct of rigorous analyses.

We add to the available body of evidence by proposing an individual-based model representative of a prototypical psychiatric-psychosomatic hospital offering treatment to patients for extended periods of time. Our study aims at providing some generalizable key results with a robust quantification of their uncertainty. This is achieved by conducting a comprehensive set of sensitivity analyses of parameters and various structural assumptions presented in an easily accessible form.

## Methods

### Simulation Structure

We simulate the propagation of infection and implementation of surveillance measures in a hospital (Oberberg Fachklinik Schwarzwald) with 60 inpatients with an average duration of stay of 8 weeks and 80 staff members. The simulated clinic is initialized with a fully susceptible population whose properties are updated daily for a total simulation time of 100 days. The facility is modelled as a semi-closed environment: Propagation of infection within the clinic is treated as a closed system, but interactions with the environment outside of the clinic can introduce infected individuals into the clinic. Figure 1A shows the different possibilities of virus intrusion into the clinic: New infectious patients may be admitted to the clinic, patients may be visited by infectious visitors or be infected on a temporary weekend leave from the clinic while staff could get infected between work shifts. Each simulation is subject to a set of parameter assumptions; the sets of parameters used in this study are extracted from the literature and summarized in Table 1.

**Table 1.** Summary of the used model parameters and their uncertainties according to literature. Upper and lower bounds are used for 1-way sensitivity analysis and they represent the existing lack of knowledge about these parameters. The term “derived from” indicates that input from the stated sources was not directly applicable in the model and required some form of subjective judgement and modification prior to the inclusion into the model.

| Name | Unit | Lower | Best | Upper | Description | Source |
| --- | --- | --- | --- | --- | --- | --- |
| Asymptomatic Fraction | [%] | 10 | 20 | 30 | Fraction of asymptomatic disease courses | [14, 30–32] |
| Asymptomatic Infectivity | [%] | 40 | 70 | 100 | Infectiousness of asymptomatic individuals compared to symptomatic individuals | [1, 14, 31] |
| False Symptoms | [agents/day] | 0.5 | 1 | 2 | Average daily amount of non-COVID-19 related symptomatic individuals | Assumed |
| False Traces | [agents/day] | 4 | 8 | 12 | Average amount of erroneously traced individuals assuming perfect tracing efficiency | Assumed |
| Heterogeneity Modifier | [1] | 2 | 4 | 6 | Scaling factor of transmission matrix in infectivity of high-risk/low-risk staff | Assumed |
| Incubation Mean | [days] | 5 | 5.5 | 6 | Mean of incubation time | [33, 34] |
| Incubation SD | [days] | 2.1 | 2.3 | 2.5 | Standard deviation of incubation time | [33] |
| Infectivity Heterogeneity | [1] | 1 | 1.5 | 1000 | Heterogeneity in individual infectivity: Shape parameter of the Gamma-distribution | Derived from [20, 21] |
| Isolation Fraction | [%] | 50 | 70 | 90 | Fraction of symptomatic individuals isolated daily | Assumed |
| Outside Infection | [1] | 0.01 | 0.04 | 0.16 | Scaling factor of infection risk outside of clinic | Assumed |
| Peak Infectiousness | [days] | -1 | 1 | 3 | Time shift of peak infectiousness relative to symptom onset | [17] |
| Prevalence | [%] | 0.005 | 0.02 | 0.08 | COVID-19 prevalence in population including non-confirmed cases | Assumed |
| R0 | [1] | 1.5 | 3 | 5 | Average number of infections an individual causes inside of clinic | Assumed |
| Symptom Mean | [days] | 3.5 | 5 | 6.5 | Mean of symptomatic infectious time | Derived from [16, 35–37] |
| Symptom SD | [days] | 1.1 | 1.5 | 1.9 | Standard deviation of symptomatic infectious time | Derived from [16, 35–37] |
| Test Compliance | [%] | 60 | 80 | 100 | Fraction of individuals compliant with repeated surveillance testing | Assumed |
| Test Sensitivity | [%] | 80 | 90 | 100 | Sensitivity of diagnostic test | Derived from [6, 29, 38] |
| Test Specificity | [%] | 98 | 99.5 | 100 | Specificity of diagnostic test | Assumed |
| Tracing Fraction | [%] | 50 | 70 | 90 | Fraction of infections reconstructed by contact tracing | Assumed |

**Figure 1.**
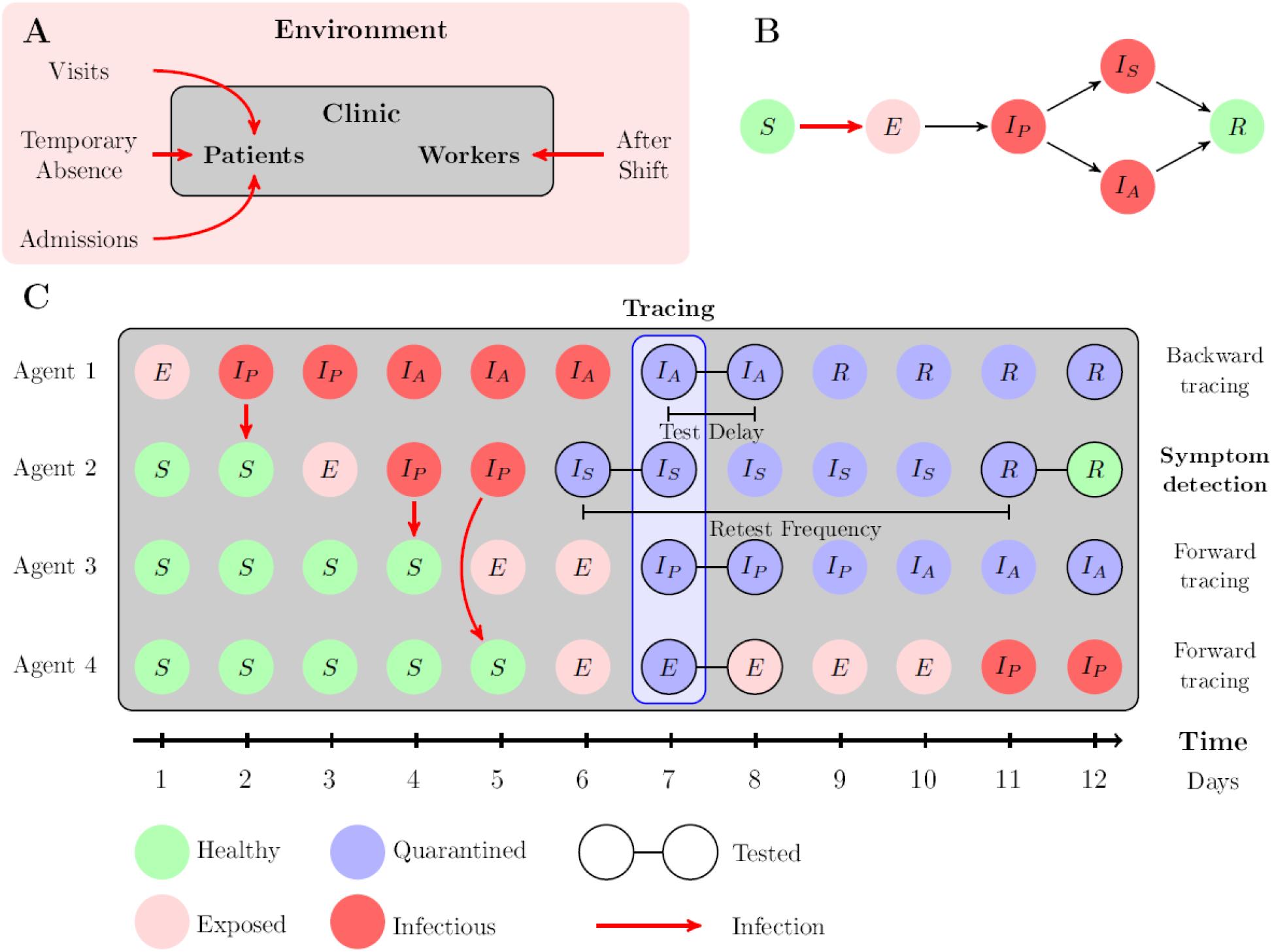
**A:** Interactions of clinic and environment. The possibilities of introducing the virus are indicated by the red arrows. **B:** General structure of disease progression. S: Susceptible, E: Exposed, I_P_: Presymptomatically Infectious, I_S_: Symptomatically Infectious, I_A_: Asysmptomatically Infectious, R: Recovered. **C:** Symptom-based baseline surveillance in a hypothetical case scenario. Agent 1 has been infected outside of the clinic (index case) and infects agent 2, who goes on to infect agent 3 and 4. On day 6, agent 2 is isolated due to developing symptoms and once the case is ascertained a day later, contact tracing isolates the primary infector (backward tracing) and subsequent infections by agent 2 (forward tracing). The isolated individuals are then tested, confirming that agent 1 and agent 3 are infectious. Agent 4 is released as the infection is not yet detectable.

### Propagation of Infection

The infection dynamics are implemented in the model at the level of individual agents which represent individuals in the clinic population. Individual-based models offer a high flexibility and allow for incorporation of inter-individual heterogeneity and inherent stochasticity, such that they are well suited to model relevant features of the epidemiological dynamics realistically [11]. The current state of disease progression is tracked for every agent individually. Based on the current state of the infected individuals within the clinic the probability of infection is derived for all susceptible individuals. The risk of infection depends on the disease states of the agents but also on various dynamic properties, such as quarantine, possible absence from the clinic and current infectivity of agents. The infection dynamics are stochastic, i.e. it is randomly drawn whether an agent is infected on a given day or not.

The disease progression is modelled structurally similar to a homogeneous stochastic SEIR-model [12], but extended to incorporate characteristic features of COVID-19 and a more realistic transmission structure. Asymptomatic individuals who display no noticeable symptoms show a significant transmission potential [13, 14]. This is partly due to presymptomatic transmission, as viral shedding begins already before symptom onset [15], i.e. during the incubation period. This leads to an extended list of states incorporated in our model: Susceptible *S*, Exposed *E*, Presymptomatically Infected *I*_*P*_, Symptomatically Infected *I*_*S*_, Asymptomatically Infected *I*_*A*_, Removed *R*. The progression of disease states is illustrated in Figure 1B.

Variations in the natural history of the disease are not only manifested in the display of symptoms but also in the timing of the different disease states. It has been shown that accounting for stochasticity in these timings significantly affects the modelled spread of a virus [12].

Consequently, the incubation time, the symptomatic time as well as the presymptomatic time are defined as random variables. The resulting distributions of these durations are visualized in Figure 2A assuming best guess parameters. The range of the uniformly distributed presymptomatic time is based on [1, 15–17], incubation time and symptomatic time are lognormal with parameters as defined in Table 1. Variations of infectiousness of individuals is driven by their different biological and behavioural components [18]. Empirically, this varying degree of infectiousness can be used to explain the *offspring distribution*, i.e. the number of secondary cases caused by the respective primary cases [19]. For transmissions of SARS-CoV-2, these offspring distributions have been observed to be highly asymmetric [20, 21], implying the existence of super-spreading. Based on these observed distributions, similar distributions have been reproduced qualitatively using our model by defining the individual infectiousness to be Gamma-distributed.

**Figure 2.**
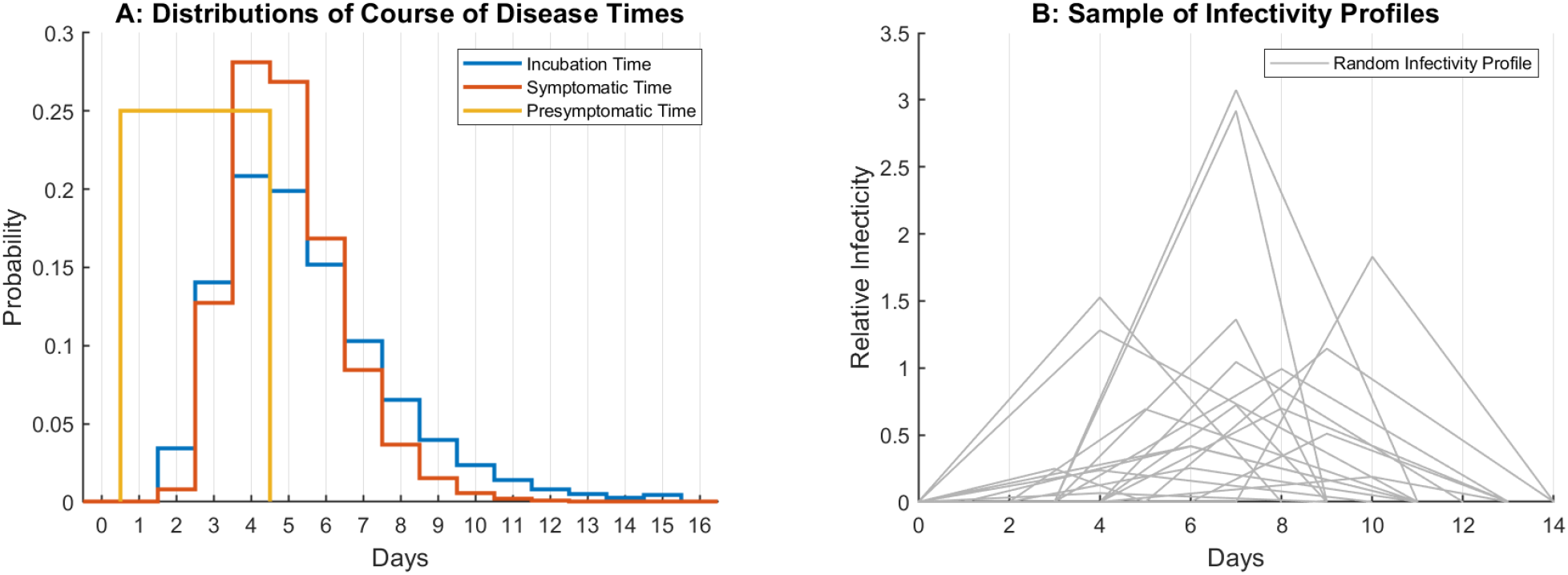
**A:** Distributions of retention times for different disease states in our simulations for the best guess parameters extracted from literature. **B:** 20 randomly drawn infectivity profiles. Each profile corresponds to the time course of infectiousness of one individual. The individual profiles have different onsets and infectivity levels.

Temporal variations of the infectiousness of agents at different stages of their disease are represented by time-dependent infectivity profiles [18, 22]. The infectivity of agents is modelled to increase linearly until a peak infectivity is reached, after which it will decrease linearly until the end of the symptomatic phase [23, 24]. Figure 2B shows a sample of infectivity profiles for 20 random individuals. The profiles vary in their behaviour over time and in their general scale, representing random time spans for disease courses and agents with different transmission potential.

Heterogeneity in the transmission structure has been found to play a key role in infection spread, e.g. by application of infection models on networks [25, 26] or age-stratified contact matrices [27, 28]. In order to implement a relevant heterogeneous transmission structure into our model, the four agent classes - *patients, low-risk staff, average-risk staff* and *high-risk staff -* are defined. The transmission rates between these classes are modified using transmission matrices, which include information about the staff’s occupation and the duration of shifts. In order to adjust for the scale of transmission dynamics, model parameters are calibrated to return pre-specified values of the reproduction number *R*_0_. A more detailed description and justification of the modelled infection dynamics and the calibration process can be found in the supplementary materials.

### Conducting surveillance

Surveillance measures are implemented to detect and prevent the spread of infection within the clinic. This is achieved by enforcing quarantine on individuals who display symptoms or individuals who were tested positive in screening measures. Once an agent is quarantined, the agent is assumed to be non-infectious during the time of isolation. The fundamental concepts of outbreak detection and containment represent a symptom-based baseline surveillance scenario which is common to all considered surveillance strategies. It comprises i) *isolation based on symptoms*, ii) *case ascertainment* via diagnostic testing, and iii) subsequent *contact tracing* to isolate contacts at high risk of possible contagion due to previous contact with the index case.

*Isolation based on symptoms* is modelled as a daily probability of isolating an agent in the symptomatic state *I*_*S*_. Typical symptoms of COVID-19 are not specific to this disease and may be mild, such that the possibility of unjustified isolations of healthy individuals is included. In order to properly deal with such ambiguous cases, a *case-ascertainment* process is needed to distinguish non-infectious from infectious individuals. Modelling case-ascertainment requires specification of a diagnostic test which can assess the disease state of an individual or, more specifically, whether the individual is infectious. The performance of the diagnostic test used in the model is uniquely specified by its sensitivity, specificity, and test-to-result delay. Performance of the PoC test used in the model is based on the Panbio™ COVID-19 antigen rapid test (Abbott) [29] as employed in the factual hospital. Finally, c*ontact tracing* disrupts possible chains of infection once infected individuals are detected. It is modelled by immediate isolation of secondarily infected individuals once the primary case has been ascertained, given a certain success probability of locating the secondary cases. Figure 1C summarizes the three mentioned concepts inherent to baseline surveillance by displaying hypothetical chains of infection and their subsequent detection via the surveillance measure.

Four different surveillance strategies are evaluated quantitatively: The aforementioned symptom-based *baseline surveillance* strategy and three *active surveillance* strategies. Active surveillance strategies comprise preventive testing of the clinic population. The first strategy is the *baseline surveillance*. The second strategy, termed *entry testing*, aims at detecting the intrusion of the virus into the clinic at the entry point, in addition to baseline surveillance. To this end, patients which are newly admitted to the clinic or who return from temporary absence are tested immediately when entering the clinic and five days after, which accounts for potentially long incubation times. The third and fourth strategy implement regular testing of the clinic population on top of entry testing and baseline surveillance, i.e. testing every agent *once weekly* or *twice weekly*, respectively. As individual agents may refuse to participate in such preventive testing measures, compliance to testing measures is defined as an additional agent property. Non-compliant agents will not be tested for strategies which impose regular testing on the clinic population, i.e. for the third and fourth strategy.

### Quantifying Outcomes

In order to quantitatively evaluate the efficacy of the different strategies, suitable outcome measures are defined. The primary outcome measure of interest is the reduction of outbreak probabilities between two defined strategies. An outbreak is defined as *N* ≥ 3 new infections over a time span of *T* = 10 days, given a 100 day simulation run. The outbreak probability is defined as the proportion of simulation runs in which an outbreak occurred. Variations of parameter assumptions affect the outbreak probability for different strategies similarly.

Consequently, ratios of outbreak probabilities of two strategies are likely less impacted by the parameter uncertainty than it is the case for absolute probability values. Stochastic uncertainty is minimized by generating at least 200.000 simulation runs for any simulation scenario, i.e. for any combination of parameters and strategy considered. The remaining stochastic uncertainty of results is indicated by error bars where applicable. In order to assess parameter uncertainty comprehensively, all model parameters are varied in a 1-way sensitivity analysis. This analysis varies one parameter at a time within its existing uncertainty, keeping all other parameters fixed at their best guess value. Parameters which have been observed to have little to no impact on results are excluded in the final analyses displayed here. In order to assess practicability of the proposed strategies, the amount of people under quarantine and the number of tests conducted per day are monitored as secondary outcomes.

## Results

### Quantifying Outbreak Probability Reduction

The relative reduction of outbreak probability by entry, once weekly and twice weekly testing relative to the symptom-based baseline strategy is displayed in Figure 3 for all parameter combinations relevant to the 1-way sensitivity analysis.

**Figure 3.**
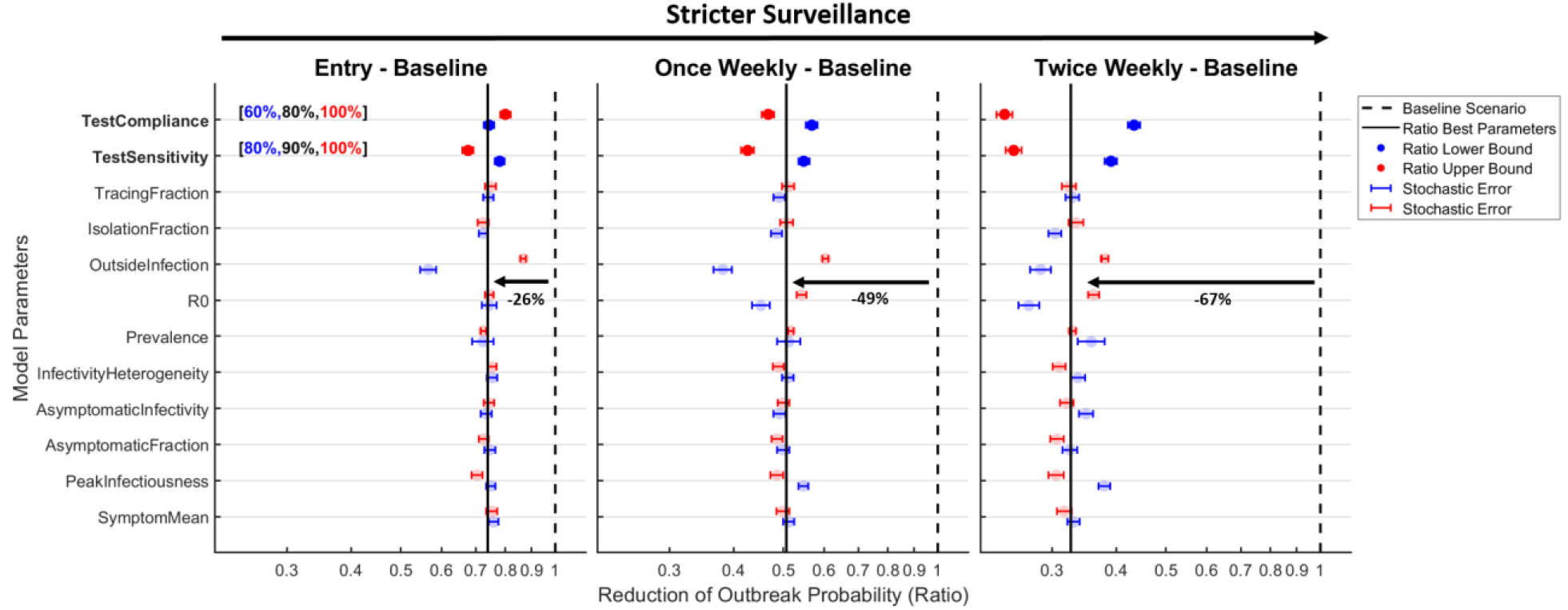
Reductions of the outbreak probabilities by entry testing, once weekly and twice weekly testing relative to the symptom-based baseline strategy on a log2-scale. The black lines correspond to the estimate of outbreak reduction for the best guess parameters. Each point corresponds to the estimated outbreak reduction for a 1-way sensitivity analysis of the corresponding parameter towards its upper bound (red) or lower bound (blue). Uncertainties due to stochasticity of the dynamics are visualized by 1σ error bars. The results for the expected reduction of the outbreak probability are robust to most epidemiological parameter assumptions, the exact parameter values employed are stated in Table 1.

Implementing entry testing reduces the probability of an outbreak by 26% relative to the baseline only strategy, additionally testing of the clinic population once or twice weekly reduces the outbreak relative to the baseline strategy by 49% and 67%, respectively. These results are indicated by solid black lines which correspond to the best guess parameters in Table 1. Deviations from best guess parameters in the 1-way sensitivity analysis are visualized by the coloured points around the black line. Blue points indicate variation of the respective parameter towards the lower bound of the uncertain parameter range, while red points indicate variation towards the upper bound. The best guess outbreak probability reductions are mostly robust to variations of parameters and the large number of simulations conducted leads to narrow stochastic uncertainties. The sensitivity of the diagnostic test and compliance of the clinic population critically determine the efficacy of the strategy. Both of these parameters are not epidemiological quantities and can therefore be targeted for optimization in applications, which due to their large impact may lead to a considerable reduction in outbreaks. Epidemiological parameters with noticeable impact on the uncertainty of results are the reproduction number (R0), the probability of contracting an infection outside of the clinic (OutsideInfection) and the timing of the peak of infectiousness (PeakInfectiousness).

In order to assess the efficacy of the symptom-based baseline surveillance, it is compared to a scenario with no surveillance in the supplementary Figure S1 analogously to the analysis in Figure 3. Conducting baseline surveillance compared to no surveillance at all reduces the probability of an outbreak by 49% assuming best guess parameters. This result lacks robustness to many parameter assumptions, ranging between values of 32% to 68%. Notably, this result is sensitive to assumptions on parameters which are important to the efficacy of symptom-based surveillance, such as the proportion of asymptomatic cases (AsymptomaticFraction), the timing of the peak of infectiousness (PeakInfectiousness), the reproduction number (R0) and the success rate of symptomatic screening (IsolationFraction).

### Test Specificity Drives Practical Feasibility

Practical feasibility of the strategies is assessed by analysing the secondary outcomes, i.e. the amount of tests conducted per day and the amount of individuals in quarantine on a given day. Since testing twice weekly is the most extensive testing strategy proposed, these outcomes have been analysed in a full 1-way sensitivity analysis. The results are visualized in the supplementary Figure S2. For the best guess parameters, approximately 37 tests are conducted daily and one individual is isolated in quarantine per day in a clinic of approximately 140 individuals. The number of tests is predominantly determined by the compliance of the clinic population. The main driver of total quarantine time is the specificity of the diagnostic test. If the specificity of the test is decreased from 99.5% to 98%, the average amount of people in quarantine per day increases 4-fold. This is reasonable since even in the absence of infections, false positives are expected regularly due to testing of the whole clinic population twice weekly. All other parameters have a comparably negligible effect, suggesting that effects such as over-sensitive symptom detection will likely not limit the practical applicability of surveillance.

### Effective Surveillance Requires Immediate Test Results

The test-to-result delay of diagnostic tests differs between PCR and PoC antigen tests. Thus, analysing the effect of this delay on the efficacy of surveillance measures may provide valuable insight regarding their usefulness in application. In order to demonstrate possible effects, the different surveillance strategies are simulated for delays of *t*_*del*_ ∈ [0,1,2] *days* in contrast to the previous analyses which assumed immediate availability of test results. Assuming previously demonstrated robustness of results, simulation runs are evaluated under the best guess parameters for all considered delays and all surveillance strategies.

Figure 4A shows the outbreak probabilities normalized relative to their value for baseline surveillance. The effect of increased surveillance is much more pronounced if the test-to-result delay is small, rendering the preventive effect of additional measures almost useless if the test-to-result delays reaches two days. The decrease in efficacy is likely due to the existence of symptomatic isolations in the baseline surveillance setting. Additional surveillance testing will only improve upon baseline surveillance if infected individuals are detected before they become symptomatic. In order to improve on a setting in which symptom-based baseline surveillance is practiced, diagnostic tests have to provide fast test results to conduct effective surveillance.

**Figure 4.**
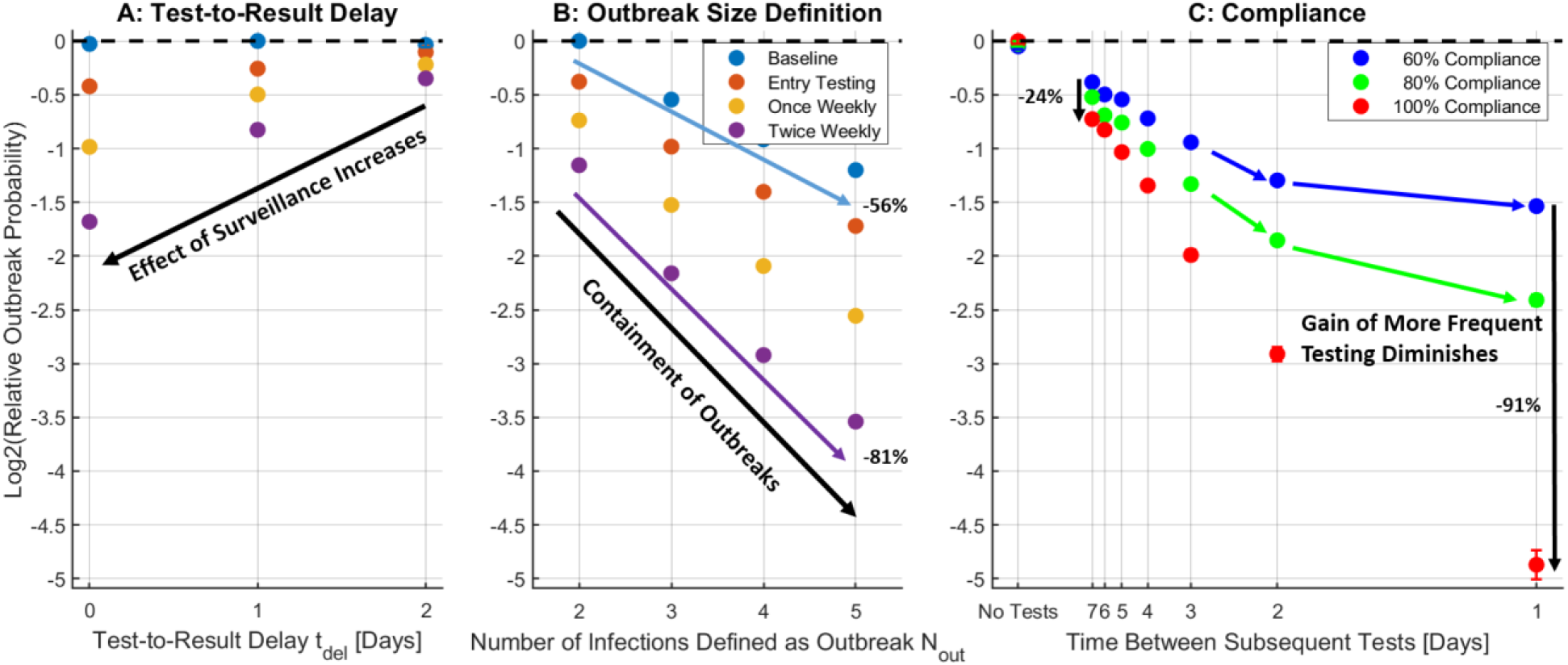
The vertical axes denote outbreak probabilities on a log 2-scale, normalized relative to the largest outbreak probability of the respective analysis. Uncertainties due to stochasticity of the dynamics are visualized by 1σ error bars, but these are mostly smaller than the point size. Results correspond to the best guess parameter set (except for changes for the particular analysis). **A:** Impact of changing the test-to-result delay on the relative outbreak probability. Decreasing test-to-result delay leads to more effective surveillance. **B:** Impact of redefining the outbreak size on the relative outbreak probability. Decreasing probabilities within a strategy implies containment of ongoing outbreaks. **C:** Impact of compliance on the outbreak probability for various regular testing frequencies implemented on top of the symptom-based baseline surveillance strategy and entry testing. The horizontal axis corresponds to a frequency scale, as test frequency is proportional to test resources required. Benefits of increasing the test frequency are limited by lack of compliance, especially if test frequency is already high.

### Strict Surveillance Improves Outbreak Containment

Variation of the outbreak size, defined earlier as ≥ 3 infected individuals over a course of 10 days, provides insight about the effective containment of outbreaks. Outbreak probabilities have been simulated for the four defined strategies and four outbreak sizes *N*_*out*_ ∈ [2,3,4,5] assuming best guess parameters. Figure 4B shows the outbreak probabilities relative to their largest value, i.e. the baseline surveillance strategy with *N*_*out*_ = 2. A decreasing outbreak probability with increasing outbreak size implies containment of occurring outbreaks. A steeper decrease is equivalent to more effective outbreak containment indicating that the more extensive surveillance strategies provide more effective containment of outbreaks. While approximately 44% of outbreaks of size 2 evolve to outbreaks of size 5 for the baseline surveillance strategy, this fraction drops to about 19% of outbreaks for testing twice weekly.

### Lack of Compliance Limits Efficacy of Regular Testing

A population fully compliant to active surveillance testing is not guaranteed in an application setting. Therefore, the impact of varying levels of compliance with regular preventive testing measures in the clinic population are investigated. Different test frequencies are analysed using the best guess parameters and assuming established baseline surveillance and entry testing. Figure 4C shows the resulting outbreak probability for different frequencies and varying levels of compliance relative to the outbreak probability of the strategy with the lowest compliance and without additional testing. Increasing compliance from 60% to 100% of the population decreases the outbreak probability by 24% if testing is conducted once weekly and by 91% if testing is conducted daily. Thus it is concluded that a lack of compliance limits the efficacy of regular testing, especially if testing frequency is high.

## Discussion

We employed an individual-based model tailored to the setting of a typical mental health treatment facility to explore different surveillance strategies intended to suppress COVID-19 outbreaks. We modelled four surveillance strategies: symptom-based baseline surveillance, entry testing, testing once a week or testing twice a week. For each active surveillance strategy, estimates for the relative reduction of outbreak probability compared to the baseline strategy were obtained. We investigated critical determinants of the epidemiological dynamics and found that fast test results, high compliance and high test sensitivity are crucial for a successful outbreak prevention. Investigation of the average number of diagnostic tests conducted and individuals under quarantine per day showed that regular testing is practically feasible if specificity of the diagnostic test is sufficiently high as this implies a low number of false positive test results.

A comprehensive sensitivity analysis confirmed the robustness of the obtained results under varying assumptions about a set of uncertain parameters. The model has been parametrized such that alternative model structures can be analysed. Examples are the parametrization of heterogeneity in the contact structure or the parametrization of different distributions of the infectiousness of individuals by scalar parameters. Moreover, the stochastic nature of the simulation model has been controlled for by generating a sufficient amount of simulation runs. Although an appropriate analysis of uncertainty has been conducted, the 1-way sensitivity analysis employed does not yield confidence or credible intervals on the generated estimates. However, the 1-way sensitivity analysis provides easily interpretable outcomes and introduces no additional bias compared to probabilistic sensitivity analyses, which require the specification of poorly known parameter probability distributions [11].

We focused on the suppression of outbreaks, defined here as 2-5 newly infected individuals over the course of 10 days. This approach differs from previous modelling studies which considered outcomes that focus on late outbreak stages, e.g. the number of cumulative infections [8–10]. The simulation of an outbreak for a long period of time amplifies the uncertainty in the assumed transmission mechanisms and in the epidemiological parameters due to the non-linear infection dynamics. Consequently, focusing the analysis on prevention of small outbreaks leads to uncertainties that can be controlled more realistically. Additionally, the small outbreak setting may reduce the impact of the modelled contact structure on the simulation results considerably. Indeed, we could see in our study that the impact of the parameter controlling the extent of heterogeneity in the contact structure was negligible. Therefore, only the variation of individual transmissibility needs to be incorporated into the model, but not an explicit contact structure as employed in many other studies [8–10, 25, 26]. Limiting results to small outbreak sizes is not restrictive in the practical application as even small outbreaks have major consequences for psychiatric clinics or skilled nursing facilities and should, therefore, be avoided as rigorously as possible.

Another important structural limitation is the assumption of constant sensitivity of diagnostic tests during the course of disease. To account for this, in the future, the course of disease could be defined based on a viral load profile across time which is proportional to infectiousness as well as the sensitivity of the diagnostic test [24]. Current evidence suggests that PoC antigen tests can indeed perform well to detect relevant levels of viral load [29], highlighting the difficulty in establishing PoC test performance when compared to PCR tests as a reference standard [5]. In order to allow for a fair comparison between surveillance based on PCR tests and PoC antigen tests different detection threshold levels for viral load have to be included in the model. However, this does not affect our conclusion that short test-to-result delays are crucial for effective surveillance.

COVID-19 surveillance in hospitals with long treatment duration and long-term care facilities provides a unique opportunity to create a safe environment for a vulnerable population. In this context, adequate assessment of the gain of various mitigation strategies is sparse but urgently needed to establish standards for practical implementation of strategies. In order to complement the existing literature, we quantified the effect of various surveillance strategies on the probability of occurring viral outbreaks. We demonstrated that implementing these strategies is practically feasible. We found that improving strategies based on isolating symptomatic individuals by means of testing-based strategies requires fast diagnostic test results, highlighting a possible use of point-of-care tests in this setting. Furthermore, we highlight the importance of a compliant population in order to maximize efficacy of regular testing. Overall, our results suggest that establishing surveillance exceeding symptom-based screening alone can successfully reduce disease burden in hospitals and long-term care facilities.

## Data Availability

MATLAB simulation code and generated data is publicly available online.

https://github.com/kreutz-lab/COVID19Surveillance

## Acknowledgements

No funding received.

## Code Availability

Simulation code and generated data is publicly available online (https://github.com/kreutz-lab/COVID19Surveillance).

## Author’s Contributions

TL and CK conceptualized the model structure. TL, CK and JT provided methodical inputs. MJM, MB and AWK informed the application setting, highlighting a relevant model scope and checked whether the model is realistic. All authors provided relevant literature and TL extracted model inputs. TL implemented the model and conducted the analyses. TL wrote the first draft and all authors revised and approved the manuscript. CK provided supervision during the whole process.

## References

1. Arons MM, Hatfield KM, Reddy SC et al. Presymptomatic SARS-CoV-2 infections and transmission in a skilled nursing facility. N Engl J Med 2020;382(22):2081–90.

2. Quicke K, Gallichote E, Sexton N et al. Longitudinal surveillance for SARS-CoV-2 RNA among asymptomatic staff in five colorado skilled nursing facilities: epidemiologic, virologic and sequence analysis. medRxiv [Preprint], 2020, doi: https://doi.org/10.1101/2020.06.08.20125989.

3. Verity R, Okell LC, Dorigatti I et al. Estimates of the severity of coronavirus disease 2019: a model-based analysis. The Lancet Infectious Diseases 2020;20(6):669–77.

4. Hao F, Tan W, Jiang L et al. Do psychiatric patients experience more psychiatric symptoms during COVID-19 pandemic and lockdown? A case-control study with service and research implications for immunopsychiatry. Brain Behav Immun 2020;87:100–6.

5. Mina MJ, Peto TE, García-Fiñana M et al. Clarifying the evidence on SARS-CoV-2 antigen rapid tests in public health responses to COVID-19. The Lancet 2021;397(10283):1425–7.

6. Guglielmi G. Rapid coronavirus tests: a guide for the perplexed. Nature 2021;590(7845):202–5.

7. Roberts M, Russell LB, Paltiel AD et al. Conceptualizing a model: a report of the ISPOR-SMDM Modeling Good Research Practices Task Force--2. Value Health 2012;15(6):804–11.

8. Smith DRM, Duval A, Pouwels KB et al. Optimizing COVID-19 surveillance in long-term care facilities: a modelling study. BMC Med 2020;18(1):386.

9. Holmdahl I, Kahn R, Hay J et al. Frequent testing and immunity-based staffing will help mitigate outbreaks in nursing home settings. medRxiv [Preprint], 2020, doi: https://doi.org/10.1101/2020.11.04.20224758.

10. Le Nguyen LK, Howick S, McLafferty D et al. Evaluating intervention strategies in controlling coronavirus disease 2019 (COVID-19) spread in care homes: An agent-based model. Infect Control Hosp Epidemiol 2020:1–11.

11. Pitman R, Fisman D, Zaric GS et al. Dynamic transmission modeling: a report of the ISPOR-SMDM Modeling Good Research Practices Task Force-5. Value Health 2012;15(6):828–34.

12. Britton T, Lindenstrand D. Epidemic modelling: aspects where stochasticity matters. Math Biosci 2009;222(2):109–16.

13. Nikolai LA, Meyer CG, Kremsner PG et al. Asymptomatic SARS coronavirus 2 infection: invisible yet invincible. Int J Infect Dis 2020;100:112–6.

14. Buitrago-Garcia D, Egli-Gany D, Counotte MJ et al. Occurrence and transmission potential of asymptomatic and presymptomatic SARS-CoV-2 infections: A living systematic review and meta-analysis. PLoS Med 2020;17(9):e1003346.

15. Wei WE, Li Z, Chiew CJ et al. Presymptomatic transmission of SARS-CoV-2 - Singapore, January 23-March 16, 2020. MMWR Morb Mortal Wkly Rep 2020;69(14):411–5.

16. He X, Lau EHY, Wu P et al. Temporal dynamics in viral shedding and transmissibility of COVID-19. Nat Med 2020;26(5):672–5.

17. Byrne AW, McEvoy D, Collins AB et al. Inferred duration of infectious period of SARS-CoV-2: rapid scoping review and analysis of available evidence for asymptomatic and symptomatic COVID-19 cases. BMJ Open 2020;10(8):e039856.

18. Grassly NC, Fraser C. Mathematical models of infectious disease transmission. Nat Rev Microbiol 2008;6(6):477–87.

19. Lloyd-Smith JO, Schreiber SJ, Kopp PE et al. Superspreading and the effect of individual variation on disease emergence. Nature 2005;438(7066):355–9.

20. Hasan A, Susanto H, Kasim MF et al. Superspreading in early transmissions of COVID-19 in Indonesia. Sci Rep 2020;10(1):22386.

21. Endo A, Abbott S, Kucharski AJ et al. Estimating the overdispersion in COVID-19 transmission using outbreak sizes outside China. Wellcome Open Research 2020;5:67.

22. Fraser C, Riley S, Anderson RM et al. Factors that make an infectious disease outbreak controllable. PNAS 2004;101(16):6146–51.

23. Chang SL, Harding N, Zachreson C et al. Modelling transmission and control of the COVID-19 pandemic in Australia. Nature Communications 2020;11:5710.

24. Larremore DB, Wilder B, Lester E et al. Test sensitivity is secondary to frequency and turnaround time for COVID-19 screening. Sci Adv 2021;7(1):eabd5393.

25. Reich O, Shalev G, Kalvari T. Modeling COVID-19 on a network: super-spreaders, testing and containment. medRxiv [Preprint], 2020, doi: https://doi.org/10.1101/2020.04.30.20081828.

26. Großmann G, Backenköhler M, Wolf V. Importance of interaction structure and stochasticity for epidemic spreading: a COVID-19 case study. In: Gribaudo M, Jansen DN, Remke A (eds). Quantitative Evaluation of Systems, 2020; vol. 12289, 211–29, Springer, Cham.

27. Wallinga J, Teunis P, Kretzschmar M. Using data on social contacts to estimate age-specific transmission parameters for respiratory-spread infectious agents. Am J Epidemiol 2006;164(10):936–44.

28. Cauchemez S, Bhattarai A, Marchbanks TL et al. Role of social networks in shaping disease transmission during a community outbreak of 2009 H1N1 pandemic influenza. PNAS 2011;108(7):2825–30.

29. Gremmels H, Winkel BMF, Schuurman R et al. Real-life validation of the Panbio™ COVID-19 antigen rapid test (Abbott) in community-dwelling subjects with symptoms of potential SARS-CoV-2 infection. EClinicalMedicine 2021;31:100677.

30. Mizumoto K, Kagaya K, Zarebski A et al. Estimating the asymptomatic proportion of coronavirus disease 2019 (COVID-19) cases on board the Diamond Princess cruise ship, Yokohama, Japan, 2020. Euro Surveill 2020;25(10):pii=2000180.

31. Lee S, Kim T, Lee E et al. Clinical course and molecular viral shedding among asymptomatic and symptomatic patients with SARS-CoV-2 infection in a community treatment center in the republic of Korea. JAMA Intern Med 2020;180(11):1447–52.

32. Rivett L, Sridhar S, Sparkes D et al. Screening of healthcare workers for SARS-CoV-2 highlights the role of asymptomatic carriage in COVID-19 transmission. Elife 2020;9:e58728.

33. Lauer SA, Grantz KH, Bi Q et al. The incubation period of coronavirus disease 2019 (COVID-19) from publicly reported confirmed cases: estimation and application. Ann Intern Med 2020;172(9):577–82.

34. Zhang P, Wang T, Xie SX. Meta-analysis of several epidemic characteristics of COVID-19. Journal of Data Science 2020;18(3):536–49.

35. Cheng H-Y, Jian S-W, Liu D-P et al. High transmissibility of COVID-19 near symptom onset. medRxiv [Preprint], 2020, doi: https://doi.org/10.1101/2020.03.18.20034561.

36. Bullard J, Dust K, Funk D et al. Predicting infectious SARS-CoV-2 from diagnostic samples. Clinical infectious diseases 2020;71(10):2663–6.

37. Wölfel R, Corman VM, Guggemos W et al. Virological assessment of hospitalized patients with COVID-2019. Nature 2020;581(7809):465–9.

38. Mohammadi A, Esmaeilzadeh E, Li Y et al. SARS-CoV-2 detection in different respiratory sites: a systematic review and meta-analysis. EBioMedicine 2020;59:102903.

