## Supplementary Material for "Preventing COVID-19 Outbreaks Through Surveillance Testing in Healthcare Facilities - A Modelling Study"

### 1 Supplementary Results

This section lists additional results which provide more insight into the results of the main text. An overview of all the conducted analyses and their properties of is provided in Table S1.

| **Analysis** | **Figure ID** | $\boldsymbol{N}_{\boldsymbol{sim}}$ | $\boldsymbol{t}_{\boldsymbol{del}}$ [d] | **Compliance** | $\boldsymbol{N}_{\boldsymbol{out}}$ | **Test Frequency** |
| --- | --- | --- | --- | --- | --- | --- |
| **4 Strategies (**$\boldsymbol{N}_{\boldsymbol{out}}\boldsymbol{=3)}$ | 3, S3 | 200.000 | 0 | [0.6,0.8,1] | 3 | [-,-,1x weekly,2x weekly] |
| **Test-Delay** | 4A | 200.000 | [0,1,2] | 0.8 | 3 | [-,-,1x weekly,2x weekly] |
| **Outbreak Size** | 4B | 2.400.000 | 0 | 0.8 | [2,3,4,5] | [-,-,1x weekly,2x weekly] |
| **Frequency/Compliance** | 4C | 200.000 | 0 | [0.6,0.8,1] | 3 | Every [1,2,3,4,5,6,7,$\infty$] days |
| **Quarantine/Tests** | S2 | 10.000 | 0 | [0.6,0.8,1] | 3 | 2x weekly |
| **Offspring Distribution** | S5 | 4000 | - | - | - | Never |
| **Baseline-No Surveillance** | S1 | 200.000 | 0 | - | [3,5] | Never |
| **4 Strategies (**$\boldsymbol{N}_{\boldsymbol{out}}\boldsymbol{=5)}$ | S4 | 200.000 | 0 | [0.6,0.8,1] | 5 | [-,-,1xweekly,2xweekly] |

**Table S1:** Brief summary of the properties of different model analyses. $N_{sim}$: Number of simulations per point in result figures, $t_{del}$: Test-to-result delay, $N_{out}$: Outbreak size definition.

#### 1.1 Comparison of Baseline Surveillance to No Surveillance

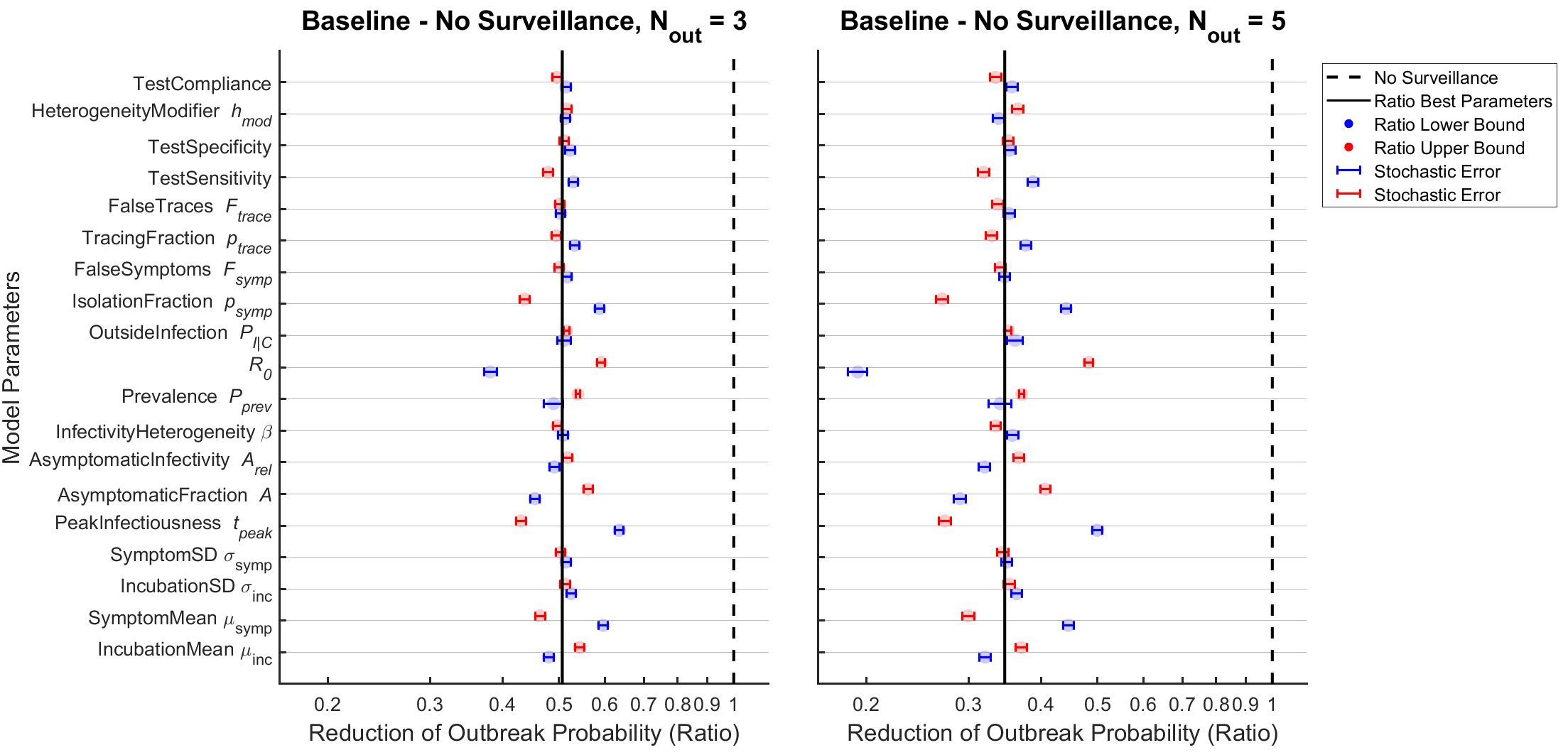

**Figure S1:** Reduction of outbreak probability for the symptom-based baseline strategy compared to no surveillance on a log2-scale for two outbreak sizes $N_{out}$. The estimate for the reduction of outbreak probability lacks robustness to many epidemiological parameters, in contrast to the main analysis in Figure 3. The effect size is sensitive to parameters which determine the efficacy of symptom-based surveillance, including the proportion of asymptomatic cases (AsymptomaticFraction), the timing of the peak of infectiousness (PeakInfectiousness), the reproduction number (R0) and the success rate of symptomatic screening (IsolationFraction). The impact of parameter uncertainty on the estimated reduction increases considerably when larger outbreak sizes are analysed.

#### 1.2 Analysis of Secondary Outcomes

***Figure S2:*** *Average number of tests conducted and average number of agents in quarantine per day for the strategy of testing twice weekly. Stochastic errors are small and can be neglected. The daily number of tests does only drastically changes with varying test compliance.* *Quarantine time is sensitive to the specificity of the diagnostic test.*

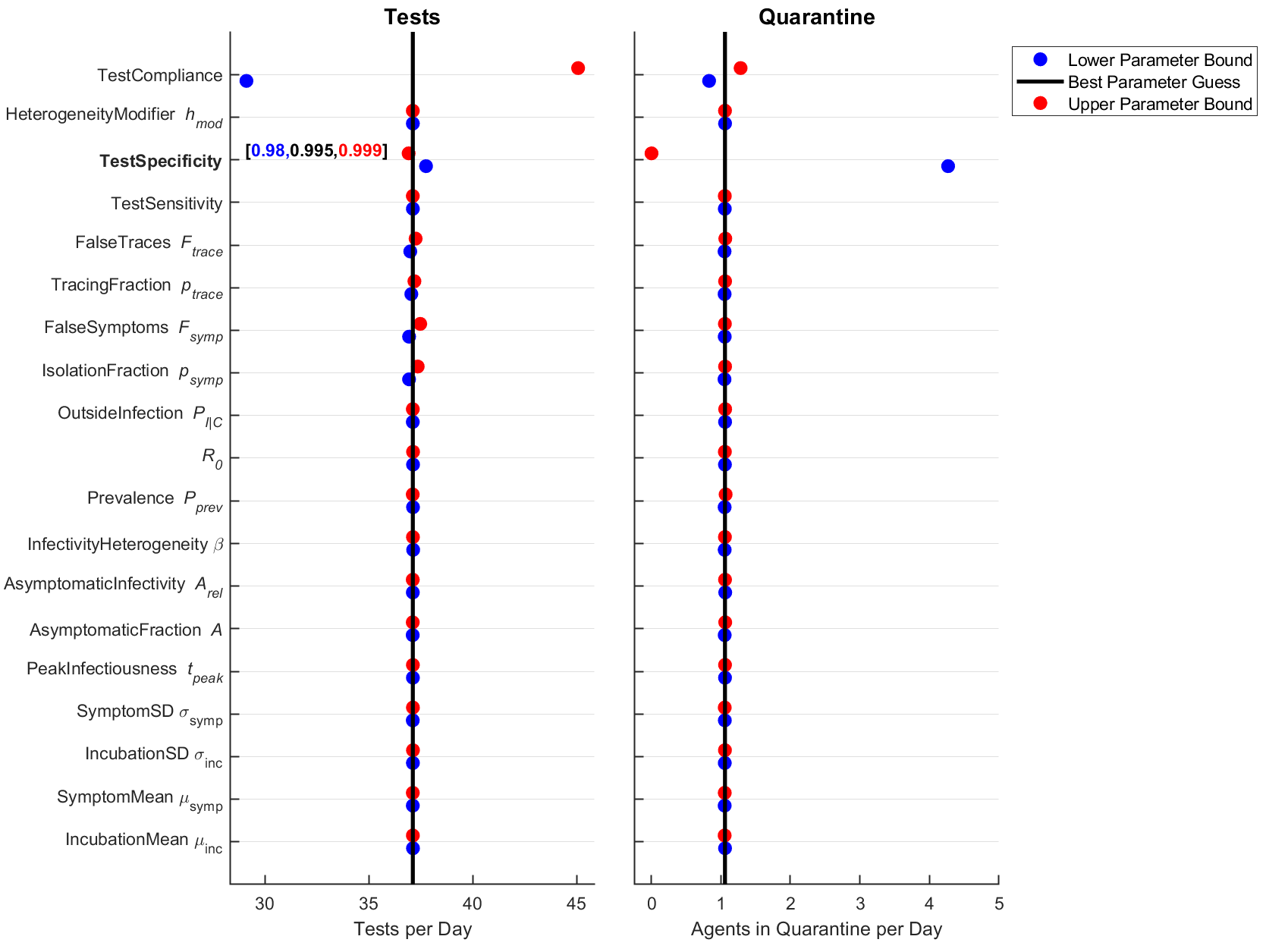

#### 1.3 Absolute Outbreak Probabilities

In the main text, the efficacy of different surveillance strategies has been quantified by relative reductions of outbreak probability between strategies rather than by absolute outbreak probability values. Figure S3 shows the absolute outbreak probabilities on which the relative reductions in Figure 3 in the main text are based on. Absolute outbreak probabilities are sensitive to parameter uncertainties, complicating the comparison between the different strategies. A strong correlation of changes in parameter and changes in outbreak probability across different scenarios is observed in this analysis. Therefore, a paired analysis considering ratios between outbreak probabilities for the same set of parameters is evaluated in the main text which is more robust against parameter uncertainties than absolute probability values.

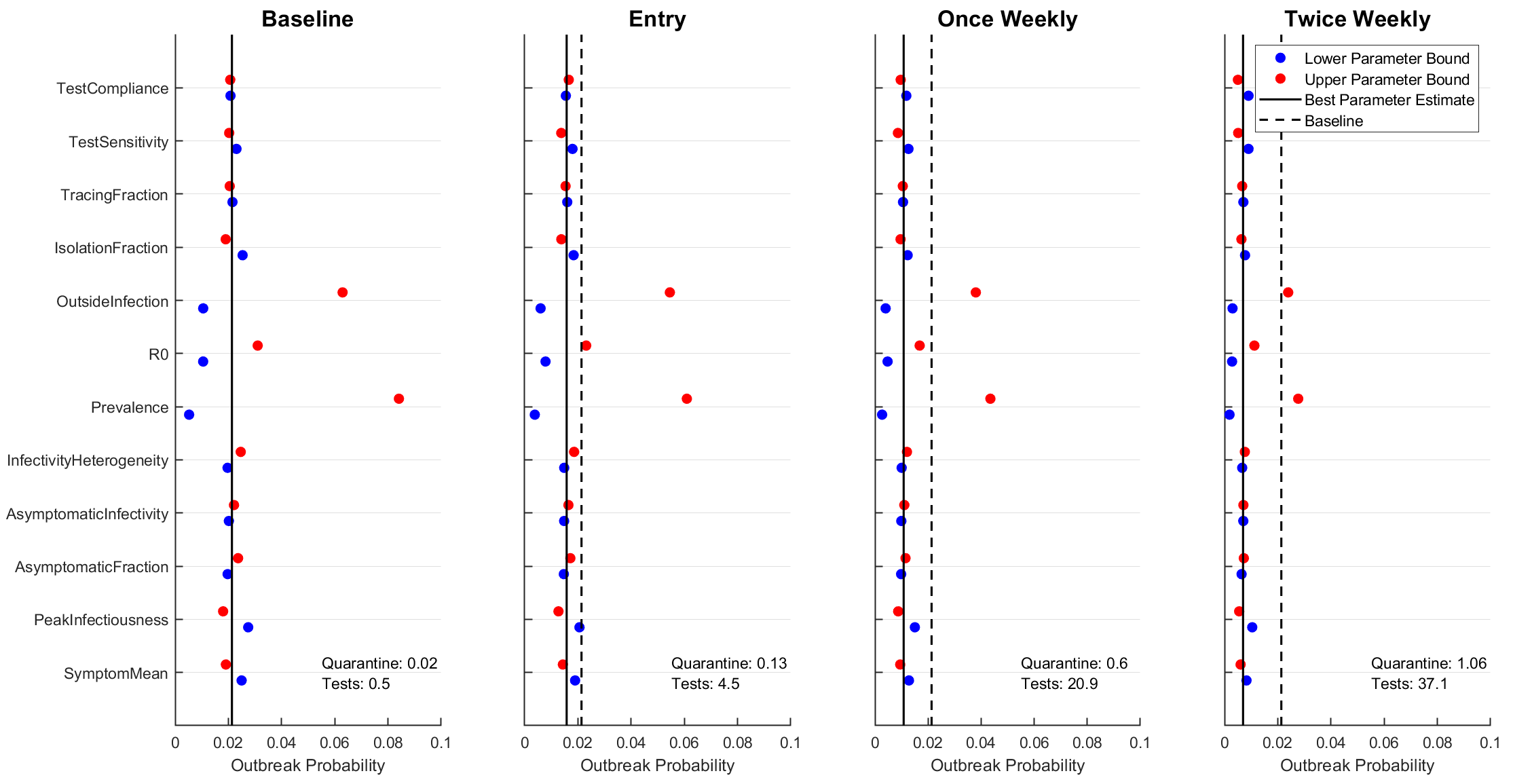

**Figure S3:** Absolute outbreak probabilities for the different strategies. Stochastic errors are smaller than the point size. The difference between the blue and red dots indicate the impact of parameter uncertainty: Absolute probabilities are sensitive to parameter variations, in particular with respect to R0, the prevalence and the probability of contact with an infected individual outside the clinic (denoted as OutsideInfection). “Quarantine” denotes the average amount of individuals under quarantine per day and “Tests” denotes the average amount of tests conducted per day.

#### 1.4 Variation of the outbreak size

An outbreak has been defined as 3 infections over the course of 10 days in most analyses in the main text. Deviation from this definition is possible by varying the amount of infections which are counted as an outbreak.

Figure S4 shows the sensitivity analysis for the comparison of the three active strategies against the symptom-based baseline strategy for an outbreak size of $N_{out}=5$. Results are less robust to parameter uncertainty as it was the case for the outbreak size $N_{out}=3$ visualized in Figure 3 in the main text. Variation of the outbreak size effectively determines for how long the infection dynamics are monitored before an outbreak is determined. Simulation of the dynamics until a large outbreak is observed amplifies the effect of existing parameter uncertainty on the outcome. This suggests that outcome measures linked to late outbreak stages might become unreliable. To further analyse the impact of uncertainty on results, consider the relative reduction of active testing compared to the baseline strategy in Figure S4 and the relative reduction of the baseline strategy to no surveillance in Figure S1. Comparing both figures reveals that the comparison of active strategy and baseline surveillance allows for conclusions much more robust to parameter uncertainties than the comparison of baseline surveillance to no surveillance.

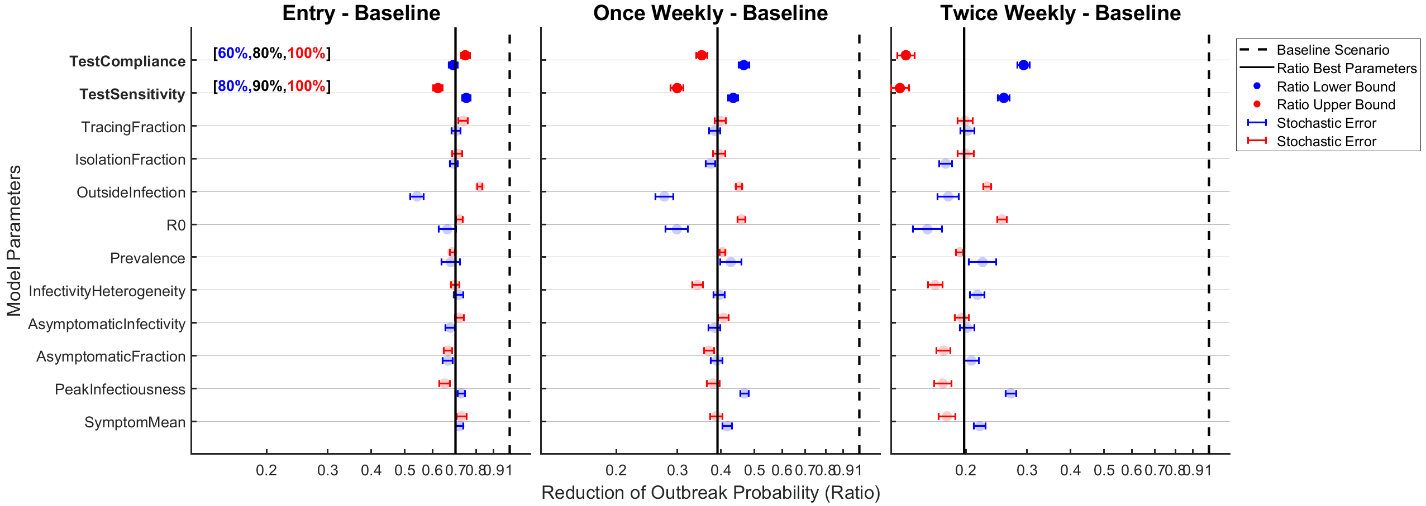

**Figure S4:** Reductions of the outbreak probabilities by entry testing, once weekly and twice weekly testing relative to the baseline strategy on a log2-scale for the outbreak size $N_{out}=5$. The impact of parameter uncertainty is more pronounced in this sensitivity analysis than it was the case for the analogous analysis for outbreak size $N_{out}=3$ in Figure 3.

#### 1.5 Efficacy of Entry Testing

The potential reduction of outbreak probability due to entry testing significantly depends on the parameter OutsideInfection as observed in the sensitivity analysis in Figure 3 and Figure S4. The parameter OutsideInfection is proportional to the probability of an agent getting infected outside of the clinic. This is also true for the prevalence, but outcomes are not sensitive to this model parameter. In order to understand this discrepancy in the efficacy of entry testing, it is helpful to subdivide the infection influx into the clinic into three contributions:

(1) Infection influx proportional to prevalence as well as OutsideInfection and reduced by entry testing (temporary leave of agents), denoted by $PO^{red}$
(2) Infection influx proportional to prevalence as well as OutsideInfection but irreducible by entry testing (staff going home between shifts), denoted by $PO^{irr}$
(3) Infection influx proportional to prevalence only and reduced by entry testing (patient admission), denoted by $P^{red}$

All three contributions to the infection influx into the clinic are proportional to the prevalence. Therefore, the relative reduction of cases due to entry testing is independent of the prevalence, as the fraction of cases detected by the measure is constant. The relative contribution of the irreducible component $PO_{irr}$ in the total infection influx is not independent of the parameter OutsideInfection: If this parameter is increased, $P^{red}$ makes up a smaller fraction of the total infection influx while the relative contribution of $PO_{irr}$ grows. As the relative contribution of the irreducible component $PO_{irr}$ to the total infection influx increases, the relative reduction of cases due to entry testing becomes smaller. This highlights the fact that the efficacy of entry testing necessarily depends on the relative contribution of cases which cannot be prevented at the entry point, i.e. by staff leaving the clinic between shifts.

### 2 Summary of Epidemiological Characteristics

#### 2.1 Incubation Time

The incubation time is defined as time from infection to symptom onset. A meta-analysis across 11 studies [1] found a mean incubation time of 5.4 days with many studies being generally consistent with this value. An estimation of incubation time from case report data including time of exposure and manifestation of symptoms reported a lognormal distribution of incubation periods with a mean of 5.5 days with a 95%-IQR of [2.2,11.5] days, corresponding to a standard deviation of 2.4 days, although case data may underrepresent mild cases [2].

The incubation time is modelled as a log-normally distributed random variable with the assumption about the distribution adopted from [2]. This distribution is determined by its mean and its standard deviation which are extracted from the reported literature. Mean incubation time analysed in the model are reasonably certain and lie in the range $\mu_{inc}\in\left[ 5,6 \right] days$ and standard deviation lies in the range of $\sigma_{inc}\in\left[ 2.1,2.5 \right] days$ based on the 95% IQR reported in [2].

#### 2.2 Symptomatic Time

The symptomatic time as defined for this study is the time from symptom onset to the end of infectiousness. Information about this period can be obtained by various methodically different means. By monitoring the transmission history of index cases to their contacts, attack rates can be stratified by time after infection, providing insight into the time-dependent infectiousness. A study of this type concerned with infections after symptom onset in Taiwan found 12 transmission pairs with 11 infections occurring in the 1-3 day span after symptom onset and just 1 the 4-5 days span [3]. Early on in the pandemic, a study of 77 non-severe transmission pairs of Guangzhou hospital admissions modelled the infectivity profile explicitly and found that the infectiousness becomes vanishingly low 8 days after symptom onset [4]. Virological studies complement the body of evidence found by such contact studies by biological examination of patient samples. Viral RNA in these samples can be detected explicitly and even investigated for their ability to reproduce, in which case the virus is likely still infectious. Detectable levels of viral RNA were reported to persist for 2 weeks and longer [5] and a scoping review reported that the time from symptom onset to two negative PCR-tests had a mean of 14 days across studies [6]. However, efforts to cultivate virus from patient samples failed for all tested samples after day 8 of symptom onset, [5, 7] suggesting that the highly sensitive detection of viral RNA is not a reliable proxy of patient infectiousness which is in accordance with reports of the scoping review [6]. Due to the technical difficulties of asserting active infectiousness, the overall evidence on the symptomatic time is scarce but suggests high infectivity around symptom onset and negligible levels of infectiousness for most individuals around day 8 after symptom onset.

The symptomatic time is modelled as a log-normally distributed random variable. Although information about the distribution could not be extracted from literature, the log-normal assumption is adopted as for the incubation time with parameters fitting the general descriptions in this paragraph. The mean symptomatic times are not entirely certain which is reflected in the large range of analysed parameters $\mu_{symp}\in\left[ 3.5,6.5 \right] days$ and the standard deviation is chosen to lie in the range of $\sigma_{inc}\in\left[ 1.1,1.9 \right] days$ to fit the available evidence.

#### 2.3 Presymptomatic Transmission

A characteristic feature of SARS-CoV-2 is a period of infectious viral shedding before symptoms manifest which is denoted as the presymptomatic time. The existence of presymptomatic transmission is reflected in a serial interval which is of a similar magnitude as the incubation time, implying that there must be significant presymptomatic transmission potential [8]. 2-point testing in a skilled nursing facility revealed that of 27 patients who were asymptomatic but SARS-CoV-2 positive at first test, 24 patients turned symptomatic with a median time of test to symptom onset time of 4 days, suggesting existence of viral RNA a couple days before symptoms appear [9]. In a study of transmission pairs, infectiousness was found to start from >2 days before symptom onset and relevant levels of infectiousness start 5 days before the onset of symptom. [4] A descriptive study of transmission clusters in Singapore found 10 presymptomatic transmissions which occurred 1-3 days before symptom onset [10]. A scoping review on infectious times found that most studies consistently report mean presymptomatic periods of 1-4 days [6].

The available evidence suggests that presymptomatic transmission is relevant and infectiousness can with some certainty be modelled to start in the range of 1-4 days before symptom onset. The presymptomatic time is modelled as a random variable which, due to lack of more specific information, is assumed to be uniformly distributed on the specified range of 1-4 days before symptom onset.

#### 2.4 Asymptomatic Transmission

Apart from the temporal properties of the natural history of disease, characteristics pertaining to asymptomatic individuals need to be included. The asymptomatic fraction, i.e. the fraction of infected cases who do not develop noticeable symptoms at any stage of the disease is of major importance when trying to predict how well different outbreak suppression strategies perform. With increasing numbers of scientific investigations in this regard, the existence of a large reservoir of completely asymptomatic individuals can be excluded. In an extensive meta-analysis, an asymptomatic fraction of 20% was reported across studies [11]. Specific studies with large numbers of participants which reported an asymptomatic fraction are the study on the Diamond Princess cruise ship [12] which reported 113 (18%) asymptomatic patients and a retrospective cohort study in Korea reporting 89 (29%) asymptomatic individuals [13]. In a screening study of healthcare workers in the UK [14], 6 completely asymptomatic individuals (11%) were identified, while this number climbed to 18 individuals (32%) if mild symptoms are counted as asymptomatic, providing a span of how symptom definition may affect the asymptomatic fraction. Although the definition of an asymptomatic case varies across studies, a general range of possible values can be specified.

In order to properly plan responses to the ongoing pandemic, the transmission potential of these asymptomatic carriers must be discussed. Cycle threshold (or $C_{t}$) values of a PCR test indicate the viral load available in the sample and they are defined as the number of reproduction cycles necessary to generate a detectable signal. Comparisons of $C_{t}$ values [9, 13] showed no significant difference in asymptomatic and symptomatic carriers. A review concerned with properties of asymptomatic carriers reported conflicting results in the literature concerning the existence of different transmission potential [15]. Another systematic review with a sample size of 5 studies found some evidence for a relative risk of infection smaller than 1 for asymptomatic carriers, but the confidence interval is consistent with 1 as well [11]. Consequently, the possibilities of asymptomatic infectiousness lower or equal to symptomatic infectiousness should both be considered.

Based on the available evidence, the fraction of cases $A$ which display no noticeable symptoms is modelled in the range of $A\in[0.1,0.3]$. Their infectiousness compared relative to symptomatic cases is denoted by $A_{rel}$ and takes values in the range of $A_{rel}\in[0.4,1]$.

#### 2.5 Reproduction Number

The most influential measure to classify the propensity of an infection to spread is the reproduction number $R_{0}$ which quantifies the average number of secondary infections emanating from a primary infection in a fully susceptible population. It is not an inherent characteristic of the pathogen but is instead highly contextual. To name a few factors, the behaviour of individuals, the current environment and protective measures like face masks or enhanced personal hygiene all impact the overall transmission rate and thus the reproduction number. To account for the highly contextual application, $R_{0}$ was varied across the wide range of $R_{0}\in[1.5,5]$ to reflect the existing uncertainty in this parameter in this specific setting.

#### 2.6 Generation Time

Assessing the effective virus propagation requires not only a reproduction number, but also the length of a typical reproduction cycle. This characteristic is covered by the generation time which is the time of the infection of a primary case to the infection of the secondary case. It is similar but not equivalent to the serial interval, which states the time difference between symptom onset of index and of secondary case. Unlike the generation time, the serial interval can also take negative values due to existence of presymptomatic transmission, but the mean can nevertheless be used as a proxy for the mean generation time. Generation times are a more fundamental measure for epidemiological characterizations, although also more complicated to infer. A study of cluster data in Singapore and in Tianjin found generation times of 5.2 days and 4.0 days respectively along with their respective distributions [16]. Serial interval estimation of publicly available data of 28 transmission pairs yielded a value of 4.7 days for the serial interval [8]. Interestingly, the study of transmission pairs in Guangzhou found that 8% of serial intervals are negative with an overall mean of 5.8 days [4]. Another study of 40 high confidence transmission pairs which directly estimated generation times found a mean generation time of 5.0 days as well as a corresponding distribution of times [17].

The generation time has not been used as a model input and can therefore be used to validate the infectivity profile to a limited extent. If infected individuals did not change their behaviour based on their disease state, the distribution of infectivity would be proportional to the distribution of generation times. Infectivity profiles can be randomly generated and thereupon generations times can be sampled to simulate the mean model generation time. This generation time estimate depends on several model parameters, i.e. the incubation time, the symptomatic time and the timing of peak infectiousness. In order to obtain the span of mean generation times which the model can realize, generation times are generated for the best parameter guess and a set of model parameters corresponding to both minimum and maximum mean generation time within the specified parameter uncertainty ranges. The minimal mean generation time realized by the model is 4.9 days, the best guess mean generation time is 6.4 days and the maximal mean generation time is 8.0 days. The stochastic error of these estimates is negligible compared to the rounding error. These generation times are on the large end compared to the generation times reported. But this is reasonably expected: Due to most individuals exhibiting symptoms at some point of their disease, they will tend to truncate the right side of their infectivity profile due to isolation. If this consideration is included in interpreting the simulation results, the generation times produced by the model fit well to the estimates in the literature.

#### 2.7 Peak Infectiousness

The time-dependent infectiousness was described in the main paper as linearly increasing at the start of the presymptomatic phase until a peak value is reached, after which it decreases linearly towards the end of the symptomatic phase. The timing of peak infectiousness has major implications for the spread of infection as it scales the infectivity of presymptomatic individuals: An early peak of infectiousness makes symptom-based isolation strategies less efficient. Peak infectivity is expected to occur around symptom onset, as the generation time is similar to the incubation time, implying common occurrence of presymptomatic transmission. This statement is supported by analysis of viral load dynamics in a scoping review of infectious periods, where viral loads were reported to peak on symptom onset or 2-4 days after [6].

The timing of peak infectiousness $t_{peak}$ is a model parameter which fixes the form of the infectivity profile. It is modelled in the range of $t_{del}\in\left[ -1,3 \right] days$ interpreted relative to the time of symptom onset. The possibility of infectiousness peaking before the onset of symptoms is therefore included.

### 3 Details of Simulation Structure

#### 3.1 Agent-based Model

Agent-based models provide features which deterministic models based on mean system behaviour such as SEIR differential equation models [18] or the Kermack-McKendrick formalism [17] cannot reproduce. In order to successfully model a system, the relevant features of its real-world counterpart need to be reflected in the model. This comprises i) the discreteness of the population as the system consists of only few agents, ii) the individual heterogeneity of these agents and iii) the inherent stochasticity of the underlying epidemiological dynamics. These aspects are generally not reproduced in deterministic approaches, but are inherent to agent-based simulation approaches. Such agent-based approaches constitute a flexible framework to model effects on the level of individual agents. Modelling on this level allows for specification of individual agent properties and explicit rules of how interaction and infection spread works in a highly customizable fashion [19]. Stochasticity of the epidemiological dynamics is readily incorporated by the stochastic simulation of the specified dynamics.

#### 3.2 Semi-Closed Environment

The infection dynamics of the system consists of two separate parts. Infection of agents outside of the clinic environment occurs by interaction with an environment with a certain number of effective contacts and some prevalence. This is in contrast to the interaction of individuals within the clinic which features a more detailed model of transmission based on the current state of infectious individuals currently present in the clinic. On each day of the simulation, agents are at risk of contracting the infection with a certain probability which is used to draw a Bernoulli random number signalling whether an infection actually occurred. If an agent becomes infected, they are assigned a random course of disease based on the epidemiological parameters discussed.

Mechanistically speaking, the probability of infection depends on the probability of contacting an infectious individual $P_{C}$ multiplied with the probability of infection transmission given an infectious contact $P_{I|C}$. These probabilities depend on agent properties, as $P_{I|C}$ is a function of the current infectivity of the corresponding agent and $P_{C}$ varies between different agents. Hence, for further discussion, these two parameters are defined as base rates and the mentioned modifications are subsequently introduced as modifiers for these base rates. In order to inform the intensity of infection spread within the clinic, the reproduction number $R_{0}$ is employed which is related to the product of these probabilities and can therefore only fix one of these two parameters. In order modify the infection risk outside of the clinic, another parameter is required in the model. When keeping $R_{0}$ fixed, the parameter $P_{I|C}$ can be used to modify the risk of infection outside of the clinic while keeping the risk of infection within the clinic fixed. The parameter $P_{I|C}$ scales the risk of infection with contacts outside of the clinic, while $R_{0}$ scales the risk of infection between clinic internal contacts.

The simulation of transmission is internally handled based on the parameters $P_{C}$ and $P_{I|C}$. Therefore, the probability of an agent being infected outside or inside the clinic is specified as a function of these parameters. This is discussed separately for the case of infection inside and outside the clinic.

#### 3.3 Infection Outside the Clinic

We define the probability of infection outside the clinic as the product

$$P_{inf}^{out}=P_{I|C}N_{c}P_{prev}$$

of the probability $P_{I|C}$ of getting infected given a contact with an infectious person, the expected number of close contacts $N_{C}$ and the local COVID-19 prevalence $P_{prev}$ in the population. The infection risk outside of the clinic is modified by the parameter $P_{I|C}$.

The expected number of close contacts has been guessed for the type of outside interaction, e.g. 0.2-1 close contacts for staff after their shift and 1-3 close contacts for patients when leaving the clinic over weekend. The exact values are generally not of critical importance, as the parameter $P_{I|C}$ (called OutsideInfection in Table 1) is explored in the range $P_{I|C}\in[0.01,0.16]$ in the conducted sensitivity analysis. However, the relative importance of different ways for the virus to intrude into the clinic affects the efficacy of entry testing, as workers after their shift are not tested contrary to patients returning from a temporary leave. This shall not be investigated further here as the exact values are hardly generalizable to other settings.

#### 3.4 Infection Inside the Clinic

##### 3.4.1 Calibrating $P_{C}$ by $R_{0}$

The reproduction number $R_{0}$ controls the intensity of infection transmission within the clinic and is treated as an external model input. Since the contact rate $P_{C}$ is required to calculate the probability of infection within the clinic, it needs to be calibrated by $R_{0}$. Employing the value for the transmission probability given a contact $P_{I|C}$, the base contact rate is calibrated to generate a pre-specified $R_{0}$. The exact equation on which the calibration is based looks as follows

$$R_{0}=P_{C}{\cdot P}_{I|C}\cdot N\cdot T\cdot(A\cdot A_{rel}+\left( 1-A \right))\cdot H$$

(1)

or, equivalently, solved for the contact probability

$$P_{C}= \frac{R_{0}}{P_{I|C}\cdot N\cdot T\cdot(A\cdot A_{rel}+\left( 1-A \right))\cdot H}$$

(2)

The variables used and the form of the equation are now subsequently explained and derived.

The probability of an infected individual transmitting the disease to any other agent in the clinic on a given day is $P_{C}P_{I|C}$, for $N$ agents in the clinic the expected number of infected individuals therefore is $R_{base}=NP_{C}P_{I|C}.$ This representation implicitly assumes that $N/{(N}-1) \approx1$ and it does not account for all modifications to infection probability, such as time-dependent infectivity profiles, less infectious asymptomatic agents and heterogeneous contact structure which are represented by $T, A, A_{rel}$ and $H$. The impact of these modifications on the reproduction number is now successively derived.

##### 3.4.2 Infectivity Profile

The time-dependent infectivity profiles of infected agents need to be accounted for when calibrating $P_{C}$. For each individual, a random *relative infectivity profile* with a peak normalized to 1 is drawn. The random *total relative infectivity* $\boldsymbol{T}$ is the sum over the relative infectivity of each day, represented by the area under the curve of the relative infectivity profile. By simulation of many values for $\boldsymbol{T}$, the expected total relative infectivity $T=E[\boldsymbol{T}$] can be estimated based on the generated sample. The reproduction number $R_{T}$ is the sum over all expected infections over the course of disease, therefore

$$R_{T}=P_{C}{\cdot P}_{I|C}\cdot N\cdot T$$

(3)

represents the reproduction number if heterogeneity in contact structure and if infectivity of asymptomatic agents is ignored.

The retention times for different stages of disease are drawn from independent distributions such that some loose constraints are needed to enforce sensible courses of disease and infectivity profiles:

- The times are drawn from discrete distributions to comply with the time-discrete model structure.
- Incubation time and symptomatic time distributions are truncated to force them onto the interval [2 days, 15 days] with the small remaining mass located on the edges of the distribution.
- If the presymptomatic time exceeds the incubation time, infectivity starts at day 1 after infection.
- The peak of infectiousness can neither occur on day 1 of infectiousness nor on the last 2 days of infectiousness.

##### 3.4.3 Asymptomatic Infectivity

Asymptomatic agents might differ in their infectivity compared to symptomatic agents. In order to account for these differences, it is noted that

$$R_{TA}=A\cdot A_{rel}\cdot R_{T}+(1-A)\cdot1\cdot R_{T}$$

(4)

represents the reproduction number which we provide as model input if heterogeneity of the transmission structure is neglected. The fraction of asymptomatic cases is denoted here by $A$ and their relative infectivity compared to symptomatic cases is denoted by $A_{rel}$. In this equation, $R_{T}$ represents the reproduction number associated with symptomatic cases.

##### 3.4.4 Heterogeneity in Transmission Structure

Heterogeneous interaction structure is introduced into the model by defining different classes of agents with varying amounts of contact between them. The four agent classes patients $P$, low-risk staff $S_{1}$, average-risk staff $S_{2}$, high-risk staff $S_{3}$ are considered as the determining factors in introducing the existing heterogeneity. The number of patients and staff is randomly drawn from the intervals $n_{pat}\in[50,60]$ or $n_{staff}\in\left[ 80,100 \right]$ and the three staff risk classes are populated with the proportions $[0.3,0.4,0.3]$.

An individual can be represented by an indicator vector ${V={(P,S}_{1},S_{2}, S_{3})}^{T}$ with zeros except for the correct identifier variable which is set to one. The intensity of transmission between two agents is scaled by two transmission matrices between these agents:

$$M_{risk}=\left( \begin{matrix} 1 & 1/{h_{mod}} & 1 & h_{mod} \\ 1/{h_{mod}} & 1 & 1 & 1 \\ 1 & 1 & 1 & 1 \\ h_{mod} & 1 & 1 & 1 \end{matrix} \right)M_{cont}=\frac{1}{2}\left( \begin{matrix} 2 & 1 & 1 & 1 \\ 1 & 1 & 1 & 1 \\ 1 & 1 & 1 & 1 \\ 1 & 1 & 1 & 1 \end{matrix} \right)$$

$M_{risk}$ describes variations in the risk of transmission due to staff occupation and has a variable parameter $h_{mod}$ (HeterogeneityModifier) which is introduced on empirical basis to investigate possible effects of heterogeneity.$M_{cont}$ describes the basic contact structure between patients and agents which reflects the limited time frame for possible patient-staff interactions compared to patient-patient interactions. The overall transmission modifier due to heterogeneous interaction between two agents is determined by the matrix

$${{(M}_{trans})}_{ij}={{(M}_{cont})}_{ij}{{(M}_{risk})}_{ij}.$$

The modification of transmission probability $c_{mod}$ from agent in state $V_{1}$ to agent in state $V_{2}$ is then given by the corresponding component of the matrix:

$$c_{mod}=V_{1}^{T}M_{trans}V_{2}$$

This modifier is multiplied with the corresponding base transmission rate to incorporate interaction heterogeneity into the model. The transmission matrix has not been informed by other sources, but sensitivity analyses of $h_{mod}$ suggests that interaction heterogeneity is of inferior importance compared to other uncertainties in the small outbreak size setting. For larger outbreak sizes, network effects are likely to be important but are not captured by the proposed model structure, hence large outbreaks were not considered as discussed in the main text.

Correcting the reproduction number equation for heterogeneity is necessary since the scale of values in the transmission matrix $M_{trans}$ is arbitrary. Thus, these values need to be normalized such that they do not change the intensity of infection transmission. The transmission matrix $M_{trans}$ determines the transmission probability between two different agents rather than two different classes. In order to quantify the number of interactions between two classes, a matrix $M_{scale}$ is defined which quantifies interaction probability on a level of classes. The composition of $M_{scale}$ is set to depend on the size of the different groups of agent classes, as, for example, there are more agents in the patient class than in the high-risk staff class. Let $f_{i}$ denote the fraction of individuals in the agent classes enumerated by $i=\left\{ 1,2,3,4 \right\}$, then define

$${{(M}_{scale})}_{ij}=f_{i}f_{j}$$

and note that $\sum_{i,j} f_{i}f_{j}=\sum_{i} f_{i}\sum_{j} f_{j}=1.$ Now define

$${{(M}_{mix})}_{ij}={{(M}_{scale})}_{ij}{{(M}_{trans})}_{ij}$$

where $M_{mix}$ is a measure which specifies how likely a transmission between two classes is. $M_{scale}$ introduces a guess about which class is likely the transmitting class, while $M_{trans}$ specifies to which class infection is likely spreading.

The matrix $M_{mix}$ is needed to define $H= \sum_{i,j} {{(M}_{mix})}_{ij}$ which is used to normalize the scale of the transmission matrix $M_{trans}$, such that it only introduces heterogeneity without changing the intensity of infection transmission. This normalization is related to the final reproduction number $R_{0}$ by virtue of

$$R_{0}=R_{TA}\cdot H$$

(5)

Combining equations (3)-(5) yield relates the reproduction number $R_{0}$ to the contact probability $P_{C}$ by virtue of equation (1), such that the the contact probability is calibrated by equation (2).

##### 3.4.5 Probability of Infection

Now that $P_{C}$ can be expressed by specification of $R_{0}$ in equation (2), the probability of infection $P_{inf}^{in}$ for an agent of class $i$ on a given day $t$ can now be specified by

$$P_{inf,i,t}^{in}=1-\prod_{j} ({1-P}_{C}\cdot\left( M_{cont} \right)_{ij}\cdot P_{I|C}\cdot\left( M_{risk} \right)_{ij}\cdot I_{t,j})$$

where the product is carried out over every infectious individual $j$ in the clinic and $I_{t,i}$ denotes the relative infectiousness on day $t$ as determined by the infectivity profile of individual $j$, including a possible decrease of infectiousness if the agent is asymptomatic.

##### 3.4.6 Empirical Offspring Distribution

Correctness of the calibration procedure can be validated internally by empirically comparing the reproduction number generated by the model with its pre-specified values. The reproduction number can be extracted from the more general concept of an offspring distribution which corresponds to a random variable describing the number of secondary infections a primary infector causes over the course of its disease.

For SARS-CoV-2, this distribution is characteristically over-dispersed. In order to model this over-dispersion, the relative infectivity of individual agents is set to a Gamma-distributed random variable with a mean value of 1 and varying shape parameters $\beta$ (InfectivityHeterogeneity). The shape parameter $\beta$ is inversely proportional to the variance $\beta\propto1/{\sigma^{2}}$when keeping the mean fixed. Effectively, the shape parameter controls the amount of over-dispersion in the offspring distribution, from equal individual infectivity for $\beta\to\infty$ to arbitrary large variance in individual infectivity for $\beta\to0$. Consequently, the structural assumption of varying individual infectivity is now parametrized by the shape parameter and can be readily included in sensitivity analyses. A visualization of the family of distributions of individual infectivity is shown in Figure S5A.

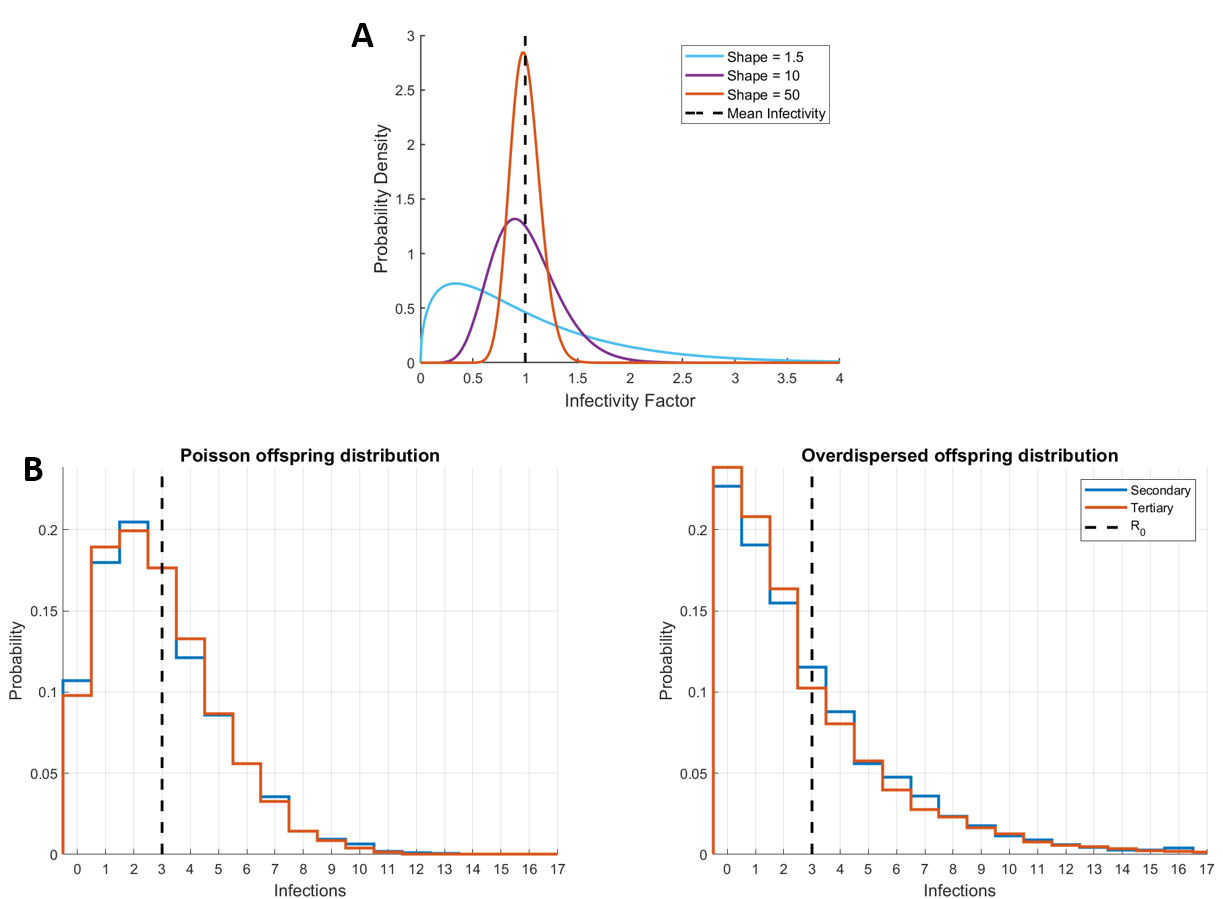

**Figure S5:** Individual infectivity and the impact on the offspring distribution. **A:** Family of gamma distributions used to sample the infectivity of individuals. The mean of each distribution is 1 and the variance is increased by modifying the shape parameter while keeping the mean constant. The shape values were chosen for illustration purposes, the values used in the model are 1, 1.5 and 1000. **B:** Offspring distributions as observed for the primary and secondary infectors in the model. If all individuals have the same infectivity, the offspring distribution is close to a Poisson distribution (left panel). Introducing individual heterogeneity with shape parameter 1.5 produces an over-dispersed distribution (right panel) which allows for more frequent occurrence of super-spreading.

Offspring distributions have been simulated by randomly seeding an infected case into the clinic and counting the number of secondary infections the index case causes. Additionally, the number of tertiary infections caused by each secondary infection is counted to assess consistency of results. There are some restrictions when counting the generated offspring to ensure that the whole course of disease is tracked which removes possibly occurring biases. Offspring distributions for homogeneous infectivity and the best guess estimate of individual infectivity are shown in Figure S5B in a simulation of unmitigated infection spread. The shape parameter of individual infectivity has been chosen to qualitatively resemble offspring distributions in empirical studies [20, 21]. It is observed that decreasing the shape parameter indeed increases the heterogeneity in the offspring distribution to a more reasonable extent. A more quantitative approach of specifying individual infectivity has been dismissed, because there are concerns with the limited amount of data and the extent to which this external data fits to the situation described by the model. However, these concerns pose no serious limitation as the uncertainty in the form of the offspring distribution is effectively addressed by sensitivity analyses.

Correctness of the model calibration is shown by the empirically generated offspring distribution in Figure S5B, as the empirical reproduction numbers of 2.92 and 2.93 for the secondary infections of both offspring distributions is close to the nominal value of $R_{0}$ = 3. This accuracy is acceptable when compared to the large range of values considered in sensitivity analyses.

#### 3.5 Simulating Surveillance

##### 3.5.1 Implementation of Surveillance model

Agent properties and dynamic states provide the necessary information which feed into the simulation framework, providing the input to simulate the model dynamics. For each simulation run, the complete dynamic history of the simulation is stored, allowing for detailed analysis of the simulation if desired. The full list of agent-specific states and properties is provided in Table S2 combined with a description of the purpose of these variables. The model dynamics are simulated as informally described in the main text. For the complete description of the model dynamics, refer to the publicly available and documented code (<https://github.com/kreutz-lab/COVID19Surveillance>).

##### 3.5.2 Baseline Surveillance

All surveillance strategies conduct symptom-based isolations as well as contact tracing which was termed baseline surveillance. Symptom-based isolation is modelled as a random daily probability $p_{symp}$ (IsolationFraction) of successful isolation of individuals who are in the symptomatic disease state. On each day, a random Bernoulli draw with success probability $p_{symp}$ is performed for every symptomatic agent to simulate whether the agent is isolated or not. In order to include individuals which show symptoms of COVID-19 but are actually not infected with SARS-CoV-2, the possibility of unjustified isolations of symptom-based screening is considered. This is implemented by randomly drawing a number of such false isolations from a binomial distribution $\mathcal{B(}N,p_{symp}\cdot F_{symp}/N)$ where $F_{symp}$ (FalseSymptoms) is the expected number of daily false isolations under perfect isolation efficiency.

Contact tracing is implemented in a similar fashion. Once contact tracing is initiated by a positive test, each infected contact is traced with a success probability $p_{trace}$ which is realized by random Bernoulli draws. In order to account for falsely traced individuals, the number of such agents is drawn from a binomial distribution $\mathcal{B(}N,p_{trace}\cdot F_{trace}/N)$ where $F_{trace}$ (FalseTraces) is the expected number of falsely traced contacts under perfect contact tracing efficiency. In both false symptomatic isolations and false contact tracing it has been implicitly assumed that the number of individuals isolated by mistake is proportional to the success probabilities of symptom-based screening and contact tracing respectively, as more strict measures should increase the number of erroneous alarms. Contact tracing is only conducted up to the first order, such that a positive test of a previously traced individual does not initiate another round of contact tracing.

##### 3.5.3 Active Strategies

Strategies involving active preventive testing of the clinic population are termed active strategies. The diagnostic test in the model is uniquely defined by its *sensitivity, specificity* and *test-to-result delay*. If an individual is in the presymptomatic, asymptomatic or symptomatic state, i.e. if the agent is infectious, the probability of a positive test result is assumed to be equal to its sensitivity. If the agent is in the susceptible, exposed or recovered state, i.e. non-infectious, the probability of a positive test result is chosen equal to 1-specificity. This implies that in our model the test accuracy is constant over the course of disease, disregarding changes in infectiousness. The test-to-result delay is defined as a lag between conducting the test and availability of its result, delaying possible countermeasures. Tested agents who are not under quarantine will isolate upon receiving a positive result, such that delays to this result lead to delays in isolation and initiation of contact tracing.

We defined the three active strategies *entry testing, once weekly* and *twice weekly* testing which define which individuals are tested on a given day. Additional analyses have been conducted to complement the results corresponding to the defined strategies, such as analysis of test-to-result delay, variation of outbreak sizes and the impact of compliance for various test frequencies. A summary of the different analyses is provided in Table S1.

#### 3.6 Number of Simulations

The number of necessary simulations depends on the outcome of interest. A low prevalence of the disease in the environment outside of the clinic leads to a small number of outbreaks within the clinic. In general, a small number of events requires a larger number of simulation runs to generate an adequate sample size to calculate the probability of an outbreak. Let $x_{out}$ be the number of observed outbreaks in a sample of $N_{sim}$ simulations and let $p_{out}$ be the true outbreak probability for the simulation scenario, then we estimate the outbreak probability and its variance by

$${\overset{^}{p}}_{out}=\frac{x_{out}}{N_{sim}}, {\overset{^}{\sigma}}_{p_{out}}^{2}=\frac{{\overset{^}{p}}_{out}(1-{\overset{^}{p}}_{out})}{N_{sim}}.$$

The values reported in Figure 4 in the main text are based on the log2-probabilities, with the maximal value being shifted to zero. The new estimate for the variance follows by Gaussian error propagation:

$${\overset{^}{\sigma}}_{\log_{2}(p_{out})}^{2}=\log_{2}(e)^{2}\frac{{\overset{^}{\sigma}}_{p_{out}}^{2}}{{\overset{^}{p}}_{out}^{2}}$$

The error bars in the main text correspond to the square root of this estimated variance.

Outbreak probabilities between two strategies are compared as a log2-ratio of these probabilities. Effects of implementing surveillance are expressed as ratios because the efficacy of strategies is expressed relative to some base case. The desired interpretation of results is therefore that decreasing the outbreak probability from 20% to 10% or from 10% to 5% corresponds to the same relative effect size. This is true on a logarithmic scale, which is why all ratios are considered on a log2-scale. Additionally, ratios on a logarithmic scale are not affected by the bound at 0 for ratios on a non-logarithmic scale and hence are usually more appropriate to describe this type of data [22]. Denote the estimated outbreak probabilities of both scenarios by ${\overset{^}{p}}_{out}, {\overset{^}{q}}_{out}$. The error for the estimate of their log2-ratio $LR$ follows by Gaussian error propagation:

$${\overset{^}{\sigma}}_{LR}^{2}=\log_{2}(e)^{2}\cdot\left( \frac{{\overset{^}{\sigma}}_{p_{out}}^{2}}{{\overset{^}{p}}_{out}^{2}}+\frac{{\overset{^}{\sigma}}_{q_{out}}^{2}}{{\overset{^}{q}}_{out}^{2}} \right)$$

Error bars in Figure 3 correspond to the square root of the variance stated here. Considering log2-ratios of outbreak probabilities between scenarios increases the amount of simulation runs required compared to stating single outbreak probabilities.

The main issue of generating an adequately sized sample of simulations is the small frequency of events, i.e. the lack of outbreaks. A small event frequency originates from the small introduction rate of infected individuals into the clinic and effective surveillance strategies which detect individuals before they cause an outbreak. All our analyses related to outbreak probability or ratios of probabilities are therefore based on a sample size of at least 200.000 simulations (see Table S1) in order to generate acceptably sized stochastic errors.

| **Name** | **Description** | **Purpose** |
| --- | --- | --- |
| ClassID | Defines agent class (patient/x-risk staff) | Calculate infection probability |
| Compliance | Binary indicator whether agents agree to regular testing | Indicates which individuals are tested in regular testing scenarios |
| DiseaseDay | Days since infection | Informs the current infectiousness and disease state |
| InfectionBy | Contains unique ID of infector if agent is infected | Information needed if contact tracing is initiated |
| InfectionCause | Source of infection (within clinic or which clinic external infection mode) | Additional analyses |
| InfectionDay | Simulation day of Infection | Reconstruction of infection dynamics |
| LastLeaveTimer | Days since agent returned to clinic after temporary leave | Indicates on which days agent is tested after return to the clinic for entry testing |
| LeaveTimer | Days since agent temporarily left the clinic | Indicates when agent returns to the clinic |
| Presence | Binary indicator whether agent is present in clinic | Indicates whether outside or inside infection scheme is used for patients |
| QuarantineTimer | Days since isolation | Indicates when agents are re-tested in isolation |
| Quarantined | Binary indicator whether agent is under quarantine | Exclude quarantined individuals as a transmission risk |
| StartDay | Simulation day of patient admission | Regulates when patient are dismissed from the clinic |
| StateID | Disease state of individual | Informs many key components of the simulation dynamics. Contains additional state for permanently dismissed patients. |
| TestDelayTimer | Days since test was conducted | If test results are not available immediately, this counter regulates when test results can be accessed |
| TestResults | Result of last test | This variable is accessed once the test-to-result delay is over and signals isolation and contact tracing if result if positive |
| Tracing | Binary indicator whether agent has been affected by contact tracing | Regulation of tracing, such that only “first-order” tracing is employed. Resets after release from isolation. |
| UniqueID | Unique Identifier of agent | Reconstruction of infection dynamics |
| Infectivity | Vector containing the relative infectivity of infected agent over time | Simulating infection dynamics |
| DiseaseCourse | Vector containing the different disease states of infected agent over time | Determines progression of disease states |

**Table S2:** Dynamic and fixed agent properties as used in the implemented simulation model. The “Purpose” section contains information about how these properties are utilized in the implemented model.

References

1.	Zhang P, Wang T, Xie SX. Meta-analysis of several epidemic characteristics of COVID-19. *Journal of Data Science* 2020;18(3):536–49.

2.	Lauer SA, Grantz KH, Bi Q et al. The incubation period of coronavirus disease 2019 (COVID-19) from publicly reported confirmed cases: estimation and application. *Ann Intern Med* 2020;172(9):577–82.

3.	Cheng H-Y, Jian S-W, Liu D-P et al. High transmissibility of COVID-19 near symptom onset. *medRxiv* [Preprint], 2020, doi: https://doi.org/10.1101/2020.03.18.20034561.

4.	He X, Lau EHY, Wu P et al. Temporal dynamics in viral shedding and transmissibility of COVID-19. *Nat Med* 2020;26(5):672–5.

5.	Wölfel R, Corman VM, Guggemos W et al. Virological assessment of hospitalized patients with COVID-2019. *Nature* 2020;581(7809):465–9.

6.	Byrne AW, McEvoy D, Collins AB et al. Inferred duration of infectious period of SARS-CoV-2: rapid scoping review and analysis of available evidence for asymptomatic and symptomatic COVID-19 cases. *BMJ Open* 2020;10(8):e039856.

7.	Bullard J, Dust K, Funk D et al. Predicting infectious SARS-CoV-2 from diagnostic samples. *Clinical infectious diseases* 2020;71(10):2663–6.

8.	Nishiura H, Linton NM, Akhmetzhanov AR. Serial interval of novel coronavirus (COVID-19) infections. *Int J Infect Dis* 2020;93:284–6.

9.	Arons MM, Hatfield KM, Reddy SC et al. Presymptomatic SARS-CoV-2 infections and transmission in a skilled nursing facility. *N Engl J Med* 2020;382(22):2081–90.

10.	Wei WE, Li Z, Chiew CJ et al. Presymptomatic transmission of SARS-CoV-2 - Singapore, January 23-March 16, 2020. *MMWR Morb Mortal Wkly Rep* 2020;69(14):411–5.

11.	Buitrago-Garcia D, Egli-Gany D, Counotte MJ et al. Occurrence and transmission potential of asymptomatic and presymptomatic SARS-CoV-2 infections: A living systematic review and meta-analysis. *PLoS Med* 2020;17(9):e1003346.

12.	Mizumoto K, Kagaya K, Zarebski A et al. Estimating the asymptomatic proportion of coronavirus disease 2019 (COVID-19) cases on board the Diamond Princess cruise ship, Yokohama, Japan, 2020. *Euro Surveill* 2020;25(10):pii=2000180.

13.	Lee S, Kim T, Lee E et al. Clinical course and molecular viral shedding among asymptomatic and symptomatic patients with SARS-CoV-2 infection in a community treatment center in the republic of Korea. *JAMA Intern Med* 2020;180(11):1447–52.

14.	Rivett L, Sridhar S, Sparkes D et al. Screening of healthcare workers for SARS-CoV-2 highlights the role of asymptomatic carriage in COVID-19 transmission. *Elife* 2020;9:e58728.

15.	Nikolai LA, Meyer CG, Kremsner PG et al. Asymptomatic SARS coronavirus 2 infection: invisible yet invincible. *Int J Infect Dis* 2020;100:112–6.

16.	Ganyani T, Kremer C, Chen D et al. Estimating the generation interval for coronavirus disease (COVID-19) based on symptom onset data, March 2020. *Euro Surveill* 2020;25(17):pii=2000257.

17.	Ferretti L, Wymant C, Kendall M et al. Quantifying SARS-CoV-2 transmission suggests epidemic control with digital contact tracing. *Science* 2020;368(6491):eabb6936.

18.	Li R, Pei S, Chen B et al. Substantial undocumented infection facilitates the rapid dissemination of novel coronavirus (SARS-CoV-2). *Science* 2020;368(6490):489–93.

19.	Pitman R, Fisman D, Zaric GS et al. Dynamic transmission modeling: a report of the ISPOR-SMDM Modeling Good Research Practices Task Force-5. *Value Health* 2012;15(6):828–34.

20.	Hasan A, Susanto H, Kasim MF et al. Superspreading in early transmissions of COVID-19 in Indonesia. *Sci Rep* 2020;10(1):22386.

21.	Endo A, Abbott S, Kucharski AJ et al. Estimating the overdispersion in COVID-19 transmission using outbreak sizes outside China. *Wellcome Open Research* 2020;5:67.

22.	Limpert E, Stahel, Werner, A., Abbt M. Log-normal Distributions across the Sciences: Keys and Clues. *BioScience* 2001;51(5):341.
